# Building resilient cervical cancer prevention through gender-neutral HPV vaccination

**DOI:** 10.1101/2023.01.17.23284655

**Authors:** Irene Man, Damien Georges, Rengaswamy Sankaranarayanan, Partha Basu, Iacopo Baussano

**Affiliations:** Early Detection, Prevention and Infections Branch, International Agency for Research on Cancer (IARC/WHO), Lyon, France; Karkinos Healthcare, Ernakulam - 682 017, India

## Abstract

The COVID-19 pandemic has disrupted HPV vaccination programmes worldwide. Using an agent-based model, EpiMetHeos, recently calibrated to Indian data, we illustrate how shifting from girls-only (GO) to gender-neutral (GN) vaccination strategy could improve the resilience of cervical cancer prevention against disruption of HPV vaccination. In the base case of 5-year disruption with no coverage, shifting from GO to GN strategy under 60% coverage (before disruption) would increase the resilience, in terms of cervical cancer cases still prevented in the disrupted birth cohorts per 100,000 girls born, by 2.8-fold from 107 to 302 cases, and by 2.2-fold from 209 to 464 cases under 90% coverage. Furthermore, shifting to GN vaccination helped in reaching the WHO elimination threshold. Under GO vaccination with 60% coverage, the age-standardised incidence rate (ASIR) of cervical cancer in India in the long-term with vaccination decreased from 11.0 to 4.7 cases per 100,000 woman-years (above threshold), as compared to 2.8 cases (below threshold) under GN with 60% coverage and 2.4 cases (below threshold) under GN with 90% coverage. In conclusion, GN HPV vaccination is an effective strategy to improve the resilience to disruption of cancer prevention programmes and to enhance the progress towards cervical cancer elimination.

## INTRODUCTION

In August 2020, the World Health Assembly adopted the Global Strategy for cervical cancer elimination, with the overarching target of reducing the age-standardised incidence rate (ASIR) of cervical cancer to fewer than four cases per 100,000 woman-years (WHO, 2020b). To reach this target, the following actions have been recommended: to fully vaccinate 90% of all girls with the HPV vaccine by age 15 years, to screen 70% of women using a high-performance test by the age of 35, and again by the age of 45, and to treat 90% of women with pre-cancer and 90% of women with invasive cancer (WHO, 2020b). However, the delivery of these interventions has been severely disrupted worldwide by the outbreak of the COVID-19 pandemic, which was declared a global emergency in the same year by the World Health Organization (WHO) (WHO, 2020a). More specifically, HPV vaccination worldwide was affected at many different levels: a) where population-based HPV vaccination programmes were already active, vaccine delivery slowed down or was interrupted (Muhoza et al., 2021), b) the launch of HPV vaccination programmes was delayed in several countries, in particular in resource-limited settings (The Lancet, 2022), and c) the production of HPV vaccines was also limited to favour the manufacturing of other vaccines (WHO, 2022a).

Apart from the COVID-19 pandemic, which has resulted in extensive disruption of health-care provision at a global level, health-care systems or public-health programmes at a local level have also regularly suffered severe disruption caused by other factors such as changes in political commitment, financial constraints, scepticism of the civil society, geo-political unrest, and environmental disasters (Colón-López et al., 2021; Gallagher et al., 2017; Germani, März, Clarinval, & Biller-Andorno, 2022; Jawad, Hone, Vamos, Cetorelli, & Millett, 2021; Larson, 2020; McGrath, 2022). Such circumstances may even lead key public health actors to reconsider their previous commitments. These partial or complete disruptions of organised disease prevention and control programmes may multiply public health crises (Sharpless, 2020), as well as provoking substantial, usually unquantified waste of human, logistic, and financial resources (Richards, Anderson, Carter, Ebert, & Mossialos, 2020). Clearly, devising and implementing pre-emptive measures aimed at improving the resilience of public health programmes would help mitigate disruption.

In a previous study, conducted before the COVID-19 pandemic in a high-income country, we showed that the addition of vaccination in boys to a programme targeting girls only could increase the resilience of the vaccination programme (Elfstrom, Lazzarato, Franceschi, Dillner, & Baussano, 2016). In the present paper, we complement our previous work by assessing the potential effect of gender-neutral HPV vaccination on resilience and progress towards the elimination of cervical cancer in India, and this in the country context where the prospect of introducing HPV vaccination into the national immunisation programme (NIP) has significantly improved following the recent marketing authorisation granted to an indigenous vaccine (The Lancet, 2022).

## MATERIALS AND METHODS

### Model

To estimate the impact of HPV vaccination on cervical cancer burden, we used the agent-based HPV transmission model, EpiMetHeos (Man, Georges, de Carvalho, et al., 2022), in combination with the cervical cancer progression model, Atlas (Bonjour et al., 2021). EpiMetHeos was used to simulate the dynamic sexual contact network through which HPV infections were transmitted and hence derive the impact of vaccination on incidence of high-risk HPV types. Based on the impact on HPV incidence, Atlas was then used to derive the impact on cervical cancer burden.

The models were previously calibrated to sexual behaviour, HPV prevalence, cervical cancer incidence in India (Man, Georges, de Carvalho, et al., 2022). Since the high-quality data on HPV prevalence and cervical cancer incidence data needed to calibrate the models to each Indian state were not available, we used a Footprinting framework to approximate the missing data and extrapolate the impact by state. In short, we first identified clusters of states with similar patterns of cervical cancer incidence. We then calibrated EpiMetHeos to the state of Tamil Nadu to represent the states in the high cancer incidence cluster, and the state of West Bengal to represent the states in the low cancer incidence cluster, using the available sexual behaviour and HPV prevalence data of these two states. Finally, we estimated the impacts for Tamil Nadu and West Bengal and extrapolated these to other states within each cluster. More details on model calibration can be found in previous publications (Man, Georges, Bonjour, & Baussano, 2022; Man, Georges, de Carvalho, et al., 2022) and in Appendices A.1-3. This study adheres to HPV-FRAME, a quality framework for modelled evaluations of HPV-related cancer control (Appendix A.4) (Canfell et al., 2019).

### Simulation scenarios

Coinciding with the recent development of a locally produced quadrivalent HPV vaccine in India (The Lancet, 2022), we simulated vaccination with a quadrivalent vaccine targeting two high-risk types, HPV16 and HPV18 (and two low-risk types, HPV6 and HPV11). As the marketing authorisation awarded to the locally produced vaccine was based on successful immune-bridging between the new vaccine and the existing quadrivalent vaccine (Gardasil™, MSD), we based our model assumptions on the efficacy estimates of the IARC India vaccine trial, which considered Gardasil (Basu et al., 2021; Joshi et al., 2022). In this trial, no difference was found between the efficacy estimates of the single-dose and two-dose schedules, which were moreover stable after up to 10 years follow-up. Hence, we based the model on pooled efficacy estimates (95% efficacy for HPV16 and HPV18, 9% cross-protection for HPV 31, HPV33, and HPV45, and 0% efficacy for the remaining high-risk HPV types) and assumed no waning of vaccine immunity over time. In doing so, the modelled efficacy could represent vaccination under either a single-dose or two-dose schedule.

Routine HPV vaccination at age 10 under the girls-only (GO) or gender-neutral (GN) strategy was modelled with different vaccination coverages in boys and girls from the range between 0% to 100%, at 10% intervals. To consider the impact of scaling up vaccination from suboptimal coverage in girls, we highlighted the following four scenarios: A) GO strategy with 60% coverage in girls, B) GO strategy with 90% coverage in girls, C) GN strategy with 60% coverage in both girls and boys, D) GN strategy with 90% coverage in both girls and boys.

To assess the resilience of different vaccination strategies on cervical cancer prevention, we simulated the above scenarios first without disruption of the HPV vaccination programme, and then with disruption occurring 10 years after the start of vaccination. As the base case, we simulated a disruption period of 5 years with total interruption of HPV vaccine delivery, i.e., 0% vaccination coverage in both boys and girls. As sensitivity analyses, we considered less strong disruption with still 20% and 40 % vaccination coverage in girls (but still 0% in boys), as well as shorter or longer durations of disruption of 1, 2, and 10 years, and no restart of vaccination. These sensitivity analyses were considered to represent different types of disruption attributable to a COVID-19 pandemic or to any other interference of the HPV vaccination programme.

### Model outcomes

Life-time number of cervical cancer cases prevented per 100,000 girls born was derived for the first 40 birth cohorts following the introduction of vaccination. Specifically, the mean number of cases still prevented across the birth cohorts with disruption, as a result of previous HPV vaccination, was defined as a measure of resilience, which was compared to the mean number of cases prevented across the birth cohorts before the disruption.

To assess progress towards cervical cancer elimination, we also derived the age-standardised incidence rate (ASIR) of cervical cancer up to 100 years after the start of vaccination. The long-term impact of vaccination was defined as the impact at 100 years after the start of vaccination. The Segi world standard population was used for the standardisation (Segi, 1960). Model-estimates of ASIR were compared to the threshold for elimination defined by the WHO of 4 cases per 100,000 woman-years (WHO, 2020b).

Model outcomes were reported as the mean and the 10th and 90th percentiles, i.e., uncertainty interval (UI), of the simulations using the 100 parameter sets best fitting the sexual behaviour and HPV prevalence data obtained through calibration.

Given the extremely low coverage of the existing screening programme in India, we have not considered the impact of screening in our study (Bruni et al., 2022).

## RESULTS

### Gender-neutral vaccination to improve resilience of cervical cancer prevention

As the base case, we simulated disruption in vaccination for 5 years, taking place 10 years after the introduction of vaccination. Life-time cervical cancer cases prevented in the birth cohorts vaccinated prior to the disruption period ranged between 562 (UI: 444, 676) and 807 (UI: 752, 853) cases per 100,000 girls born in the four highlighted scenarios. The highest impact was achieved by the GN strategy with 90% coverage, closely followed by GO with 90% coverage, then GN with 60% coverage, and then GO with 60% coverage (Figure 1).

**Figure 1.**
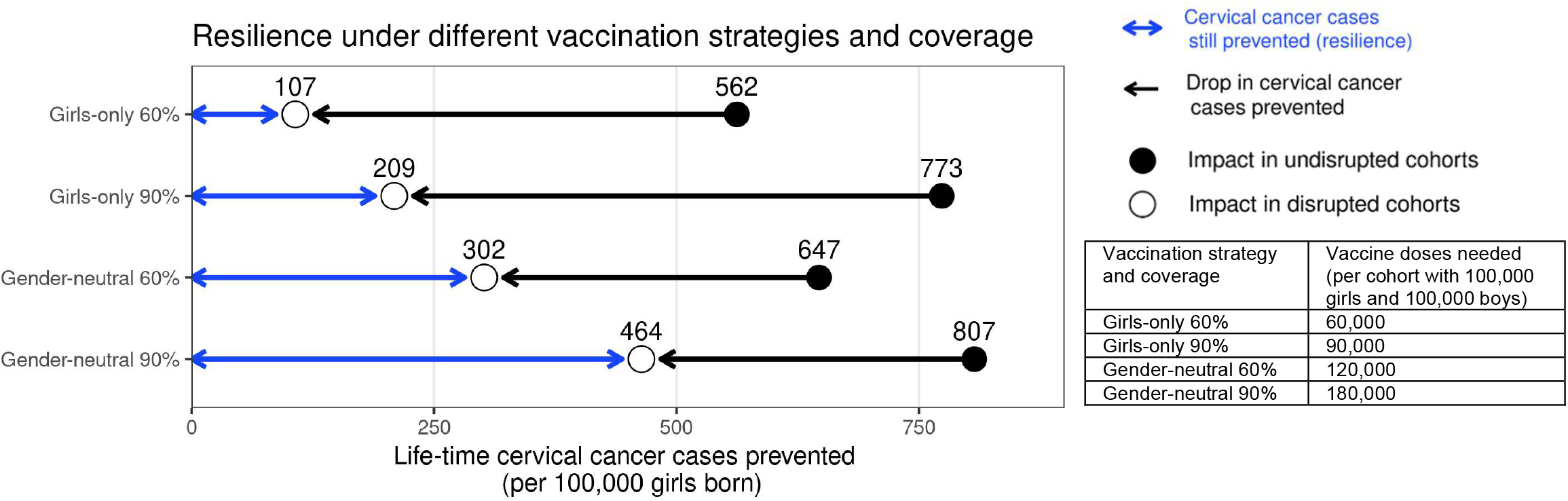
Resilience against HPV vaccination disruption in the base case. Predicted resilience, defined as life-time number of cervical cancer cases still prevented in the birth cohorts with disruption of vaccination per 100,000 girls born (blue arrow), and drop in cervical cancers prevented as compared to impact in the cohorts vaccinated prior to the disruption (black arrow), under the four highlighted scenarios. Disruption was simulated according to the base case with a period of disruption of 5 years and 0% coverage in girls and boys during the disruption period.

Among the four highlighted scenarios, the GO strategy with 60% coverage was also the least resilient (Figure 1). We found that the life-time number of cervical cancer cases prevented would drop considerably from 562 (UI: 444, 676) cases in the undisrupted cohorts to 107 (UI: 7, 214) cases per 100,000 girls born in the disrupted cohorts (i.e., resilience of 107). Increasing the coverage under the GO strategy from 60% to 90% would lead to a small absolute gain in resilience, from 107 (UI: 7, 214) to 209 (UI: 81, 340) cases prevented per 100,000 girls born, and this despite the larger increase in impact in the undisrupted cohorts vaccinated prior to the disruption from 562 (UI: 444, 676) to 773 (UI: 701, 836) cases prevented. By contrast, switching from GO to GN vaccination, under 60% coverage, led to a moderate gain of impact in the undisrupted cohorts from 562 (UI: 444, 676) to 647 (UI: 539, 746) cases prevented, but a substantial increase in resilience from 107 (UI: 7, 214) to 302 (UI: 170, 437) cases prevented per 100,000 girls born. Finally, GN vaccination with 90% coverage led to the highest impact of 807 (UI: 752, 853) cases prevented per 100,000 girls born in the undisrupted cohorts as well as the highest resilience of 464 (UI: 328, 602) cases prevented per 100,000 girls born in the disrupted cohorts.

Considering the required number of vaccine doses, we found a trade-off between dose-efficiency and resilience. In general, GO vaccination favoured dose-efficiency and GN vaccination favoured resilience (Figure 1). For instance, increasing GO coverage from 60% to 90% would lead to a higher impact in undisrupted cohorts, while needing less additional vaccine doses than switching from GO to GN vaccination under 60% coverage. However, this switch from GO to GN vaccination would yield a higher gain in resilience than when increasing GO coverage from 60% to 90% (2.8-fold versus 2-fold increase). Similarly, switching from GO to GN vaccination under 90% coverage would require an additional doubling of the number of doses and only marginally improve the impact in the undisrupted cohorts, but would produce a 2.2-fold gain in resilience.

The same trade-off between dose-efficiency and resilience was found when stratifying by Indian state. Interestingly, in the states with high cervical cancer incidence, we found slightly lower resilience under the GO strategy and a slightly greater relative gain in resilience when switching from GO to GN vaccination than in states with low incidence (supplementary Table B1, supplementary Figures B1-3). In the base case, for example, the resilience ratio for switching from GO to GN strategy (under 60% coverage) was 3-fold in states with high cervical cancer incidence and 2.7-fold in states with low cervical cancer incidence.

Sensitivity analyses on less strong disruption in vaccination coverage or alternative durations of disruption also yielded higher resilience with GN than GO vaccination (Table 1, supplementary Figures B2-3). The gain in resilience by switching from GO to GN vaccination was most evident under complete disruption of vaccination (i.e., 0% coverage at disruption) but still consistently found under less strong disruption (i.e., 20% or 40% coverage at disruption) (Table 1 part II, supplementary Figure B2). As expected, resilience decreased with longer duration of disruption, with any vaccination strategy, but was always higher with the GN strategy (Figure 2).

**Table 1.** Sensitivity analyses on coverage at disruption and duration of disruption on resilience. Life-time number of cervical cancer cases prevented per 100,000 girls born in birth cohorts vaccinated prior to disruption in part I. Sensitivity analyses on coverage at disruption in part II and on duration of disruption in part III on resilience (defined as the life-time number of cervical cancer cases still prevented in the birth cohorts with disruption of vaccination per 100,000 girls born) and resilience ratio (defined as fold change in resilience by switching from one scenario to another). Uncertainty intervals are reported in brackets.

*I. Life-time number of cervical cancer cases prevented prior to disruption*
| Scenario | GO 60% | GO 90% | GN 60% | GN 90% |
| --- | --- | --- | --- | --- |
| No disruption | 562 (444, 676) | 773 (701, 836) | 647 (539, 746) | 807 (752, 853) |

*II. Sensitivity analyses on coverage at disruption (with duration of disruption fixed at 5 years)*
| Coverage at disruption in % | Resilience by vaccination strategy and coverage |  |  |  | Resilience ratio |  |  |
| --- | --- | --- | --- | --- | --- | --- | --- |
|  | GO 60% | GO 90% | GN 60% | GN 90% | GO 60% to GO 90% | GO 60% to GN 60% | GO 90% to GN 90% |
| 0 (base case) | 107 (7, 214) | 209 (81, 340) | 302 (170, 437) | 464 (328, 602) | 2.0 | 2.8 | 2.2 |
| 20 | 271 (155, 391) | 355 (221, 490) | 425 (297, 559) | 550 (416, 680) | 1.3 | 1.6 | 1.6 |
| 40 | 410 (277, 534) | 476 (343, 599) | 527 (401, 647) | 621 (500, 730) | 1.2 | 1.3 | 1.3 |

*III. Sensitivity analyses on duration of disruption (with coverage at disruption fixed at 0%)*
| Duration of disruption in years | Resilience by vaccination strategy and coverage |  |  |  | Resilience ratio |  |  |
| --- | --- | --- | --- | --- | --- | --- | --- |
|  | GO 60% | GO 90% | GN 60% | GN 90% | GO 60% to GO 90% | GO 60% to GN 60% | GO 90% to GN 90% |
| 1 | 137 (26, 253) | 261 (125, 407) | 365 (215, 502) | 517 (372, 655) | 1.9 | 2.7 | 2.0 |
| 2 | 125 (17, 233) | 240 (105, 375) | 344 (206, 480) | 500 (359, 642) | 1.9 | 2.7 | 2.1 |
| 5 (base case) | 107 (7, 214) | 209 (81, 340) | 302 (170, 437) | 464 (328, 602) | 2.0 | 2.8 | 2.2 |
| 10 | 80 (0, 182) | 154 (33, 275) | 226 (96, 358) | 382 (240, 525) | 1.9 | 2.8 | 2.5 |

**Figure 2.**
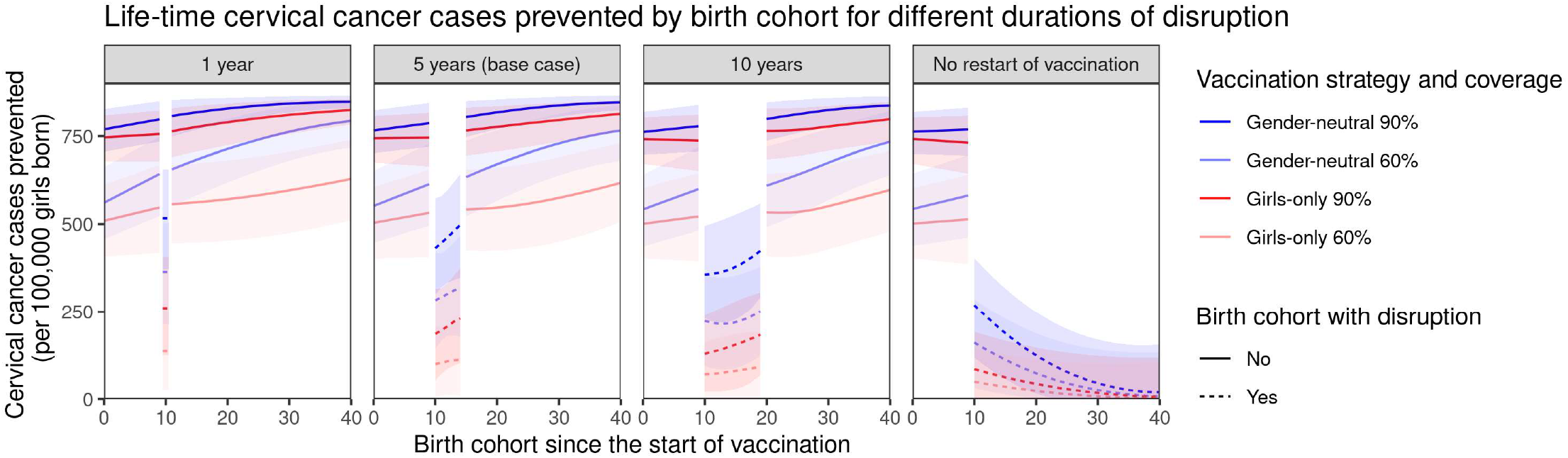
Resilience against HPV vaccination disruption in sensitivity analyses on duration of disruption. Predicted life-time cervical cancer cases prevented by birth cohorts (per 100,000 girls born) under different durations of disruption (panel) and in the four highlighted scenarios: girls-only strategy with 60% coverage (light red), girls-only strategy with 90% coverage (dark red), gender-neutral strategy with 60% coverage (light blue), and gender-neutral strategy with 90% coverage (dark blue). Vaccination coverage was fixed at 0% in girls and boys during the period of disruption. Birth cohort 0 corresponds to the first vaccinated cohort. Birth cohorts with disruption correspond to dashed lines and birth cohorts without disruption to solid lines.

The gain in resilience by switching from GO to GN vaccination was consistently above 2.7-fold under 60% coverage and above 2.0-fold under 90% coverage (Table 1 part III). Finally, in the scenario with no restart of vaccination, GN vaccination could ensure some resilience of at least 125 cases prevented per 100,000 girls born for eight birth cohorts from the start of disruption (Figure 2).

### Gender-neutral vaccination to enhance progress towards elimination of cervical cancer

Subsequently, we assessed the feasibility of reaching the WHO elimination threshold for ASIR of cervical cancer under highlighted scenarios. Introducing GO vaccination with 60% coverage would have the lowest impact, reducing the nationwide ASIR of cervical cancer from 11 cases before the introduction of vaccination to 4.7 (UI: 4.3, 5.1) cases per 100,000 woman-years in 100 years, but this remained above the WHO elimination threshold (Figure 3). The other highlighted scenarios enabled elimination to be reached within 55 to 62 years. Introducing GN vaccination with 60% coverage in both girls and boys would reduce incidence down to 2.8 (UI: 2.5, 3.2) cases per 100,000 woman-years, hence below the elimination threshold. The GO strategy with 90% coverage, as recommended by WHO, would also decrease incidence below the threshold to 2.4 (UI: 2.3, 2.6) cases per 100,000 woman-years. Lastly, given the already high vaccination coverage of 90% in girls, switching to the GN strategy would only marginally reduce incidence to 2.3 (UI: 2.2, 2.3) cases per 100,000 woman-years.

**Figure 3.**
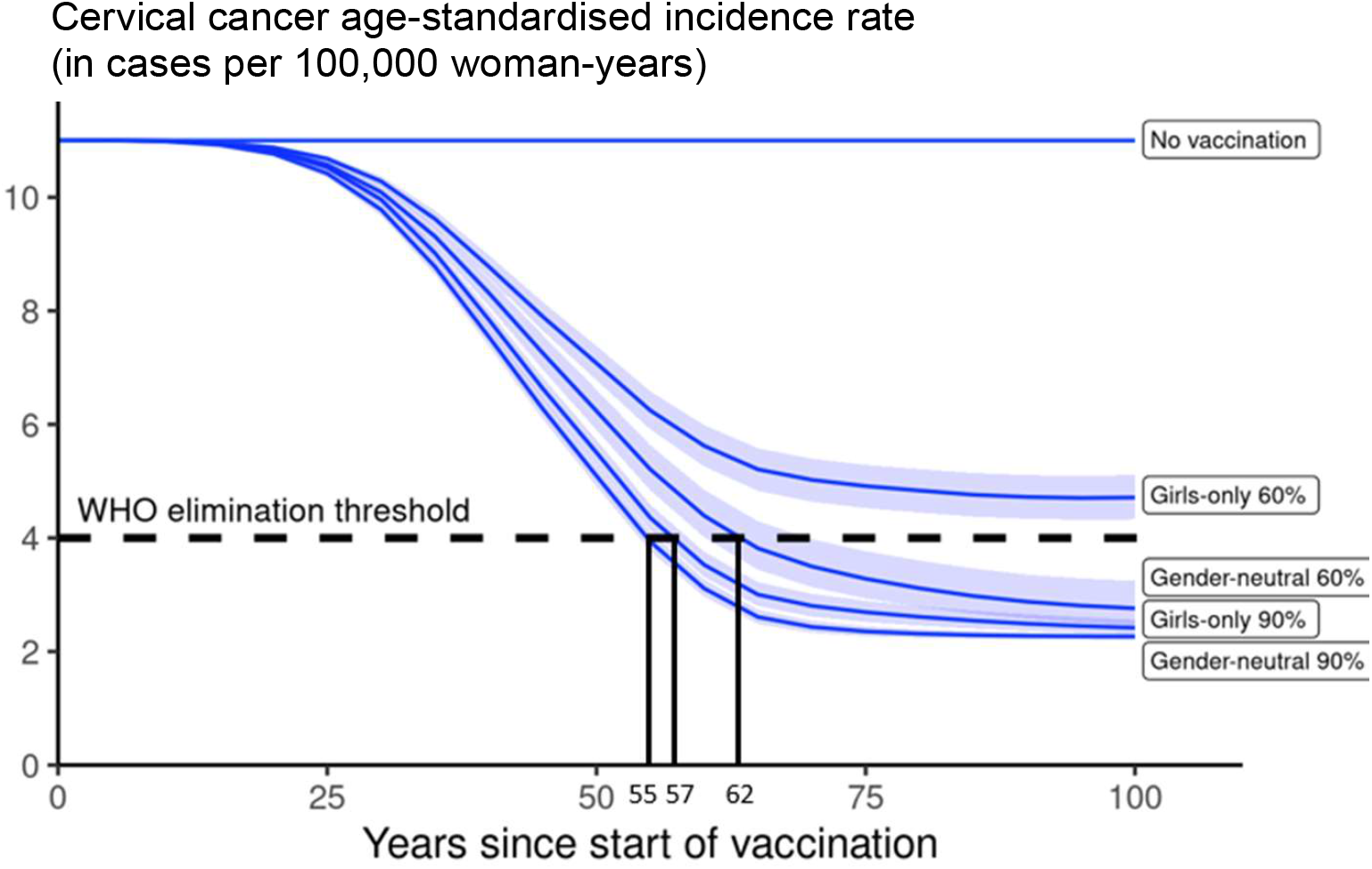
Progress towards cervical cancer elimination over time. Predicted cervical cancer age-standardised incidence (in cases per 100,000 woman-years) in the years since start of vaccination in India under no vaccination and in the four highlighted scenarios. The dashed line represents the WHO elimination threshold for cervical cancer elimination as a public health priority, i.e., age-standardised incidence of 4 cases per 100,000 woman-years. Disruption was simulated according to the base case with a period of disruption of 5 years and 0% coverage in girls and boys during the disruption period.

Other combinations of coverage in boys and girls would also allow elimination. Without vaccination in boys, the WHO elimination threshold could be reached through a critical GO coverage of 70% (UI: 65, 73) (Figure 4). With vaccination in boys, a lower critical coverage in girls might be sufficient for elimination. For instance, coverage between 50% and 70% in girls might also be sufficient for elimination when combined with moderate coverage (30%) in boys.

**Figure 4.**
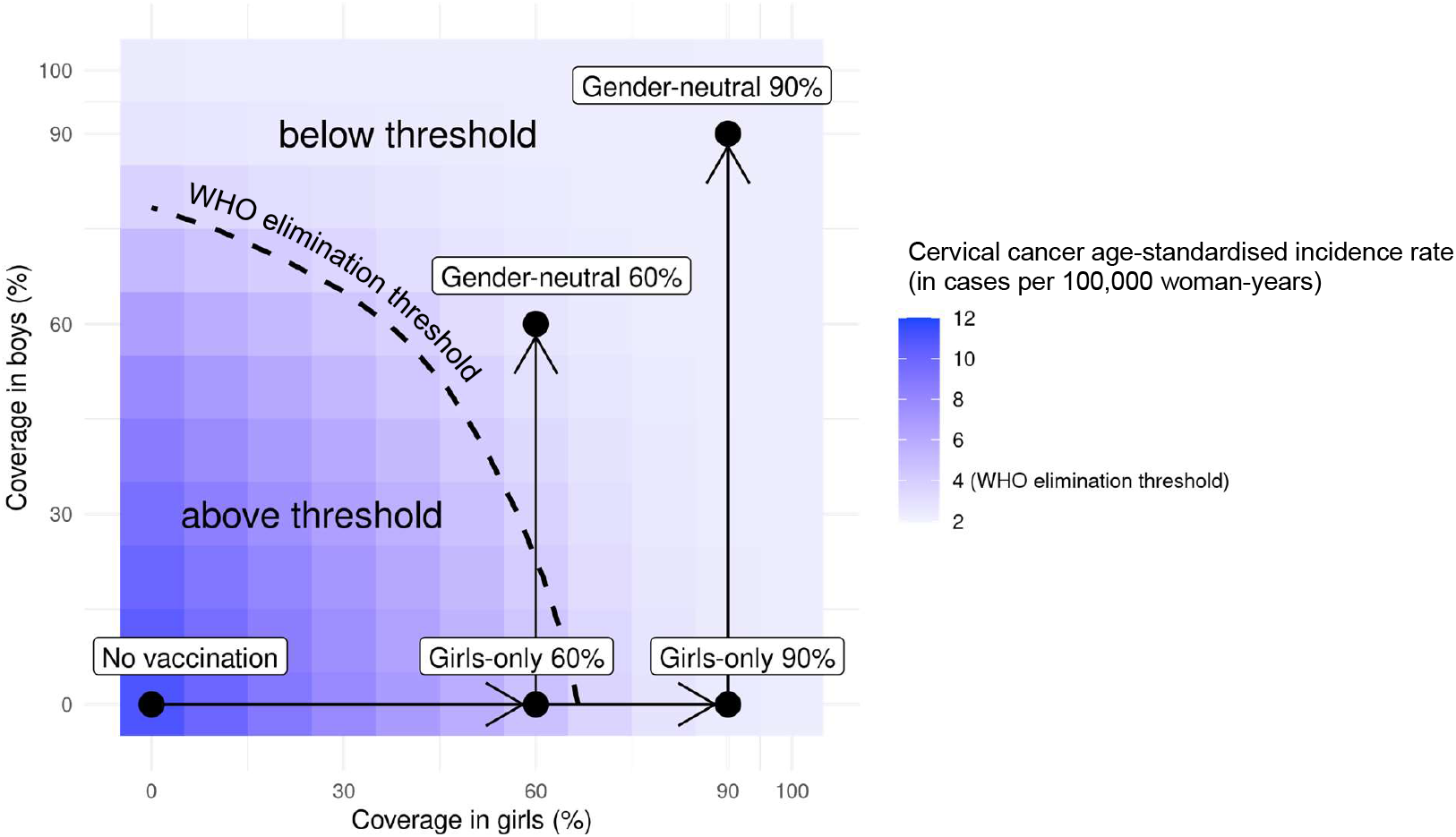
Attainment of cervical cancer elimination in the long term. Heatmap of the predicted cervical cancer age-standardised incidence rate (in cases per 100,000 woman-years) in the long-term (i.e., at 100 years after the start of vaccination) in India by vaccination coverage in girls and boys (under no disruption of vaccination). The dashed curve represents the WHO elimination threshold for cervical cancer elimination, i.e., age-standardised incidence of 4 cases per 100,000 woman-years. The five black circles correspond to no vaccination and the four highlighted scenarios: girls-only strategy with 60% coverage, girls-only strategy with 90% coverage, gender-neutral strategy with 60% coverage, and gender-neutral strategy with 90% coverage. The horizontal arrows represent scale-up of vaccination by increasing coverage in girls, and the vertical arrows represent switching to gender-neutral vaccination.

Finally, while disruption of vaccination does not prevent countries from reaching the elimination threshold eventually (provided vaccination resumes after disruption), it can prolong the number of years taken to attain this goal (supplementary Figure B4). Under the GO strategy with 90% coverage, for instance, disruption of vaccination for 5 years (as in the base case) would delay reaching elimination from 57 to 65 years since the start of vaccination. By switching to GN strategy (under 90% coverage), elimination could be reached in 58 years since the start of vaccination, even with disruption.

## DISCUSSION

As observed worldwide during the COVID-19 pandemic, public health programmes can be dramatically impacted by the occurrence of sudden societal and infrastructural crises (WHO, 2021). The repercussions of disruption of healthcare systems may subsequently translate into increased morbidity and mortality (Sharpless, 2020), and economic burden (Richards et al., 2020), at a population-level. In the present paper, using India as an example, we illustrate how the introduction of GN HPV vaccination could mitigate the negative impact of a sudden HPV vaccine delivery disruption. More specifically, using a validated agent-based model, calibrated to context-specific data, we have simulated a range of plausible scenarios of disruption of a national HPV vaccination programme and assessed the gain in resilience by shifting from GO to GN vaccination in terms of the number of cervical cancer cases still prevented in the birth cohort with vaccination disruption.

Our model-based exercise showed that in the case of HPV vaccination disruption for 5 years (base case), resilience increased by 2.8-fold (from 107 to 302 cases per 100,000 girls born) and 2.2-fold (from 209 to 464 cases per 100,000 girls born) when shifting from GO to GN vaccination under 60% and 90% coverage, respectively. GN vaccination was shown to improve resilience irrespective of the duration of disruption. However, the absolute magnitude of resilience and of resilience gain resulting from adding boys to routine vaccination of girls steadily decreased with increasing duration of disruption, under all the vaccination strategies assessed, indicating that measures to restore HPV vaccination should be implemented as soon as feasible.

The main determinant underlying the gain in resilience by switching from GO to GN vaccination is the age difference between sexual partners, with men being on average older than women within sexual partnerships almost everywhere worldwide (Wellings et al., 2006). In cases of vaccination disruption, the birth cohorts of boys vaccinated before the disruption would indirectly protect the cohorts of younger girls who missed out on vaccination during the disruption period. Since sexual behaviour is regulated by population-specific social norms, the magnitude of the resilience attributable to GN HPV vaccination is also expected to be different across populations. In India, for example, the age difference between male and female partners is on average 7 years (USAID, 2017), similar to the number of birth cohorts still retaining some protection by GN vaccination when we simulated no restart of vaccination after disruption.

In the present paper, we have also illustrated how the introduction of GN vaccination across India would enhance progress towards the elimination of cervical cancer. In principle, elimination across India could be reached with a critical level of GO coverage of 70%, which falls between the average coverage observed among girls aged 9 to 15 years living in countries with an active HPV vaccination programme, i.e., 53% (Bruni et al., 2021), and the coverage recommended by the WHO, i.e., 90% (WHO, 2020b). On the other hand, our model-based estimates suggest that cervical cancer elimination could be reached with 50% to 70% coverage in girls when combined with moderate coverage in boys, even if coverage in boys were lower than that of girls. In cases where GO coverage is already above 70%, GN vaccination would allow even more ambitious cervical cancer control targets to be reached, which might eventually make cervical screening targeted to vaccinated birth cohorts redundant (Tota, Isidean, & Franco, 2020). Moreover, as previously reported (Man, Georges, de Carvalho, et al., 2022), reaching elimination might be challenging in the Indian states with the highest baseline cervical cancer incidence before vaccination. In these cases, additional measures such as GN vaccination would be even more relevant.

A limitation of our study is that the critical levels of HPV vaccination coverage found for India are not necessarily generalisable to other populations. Baseline cervical cancer incidence and the magnitude of herd effect, i.e., the indirect protection offered by the vaccinated to the unvaccinated individuals of a population against HPV infection (Malagón, Laurie, & Franco, 2018) should be considered. These are, in turn, governed by local sexual behaviour (Bosch et al., 1994; Schulte-Frohlinde, Georges, Clifford, & Baussano, 2021). As mentioned above for resilience, the critical level of vaccination coverage to reach cervical cancer elimination will be context-specific (Baussano, Lazzarato, Ronco, & Franceschi, 2018; Lehtinen, Gray, Louvanto, & Vänskä, 2022). Extrapolation of the results of this study to other populations will be limited to those sharing a similar set of local social norms to India. Nevertheless, we expect the principle of improving the resilience of cervical cancer prevention through a shift to GN vaccination to be widely applicable, and this is supported by the consistency of our results in India when stratifying by state-specific cervical cancer incidence.

Another limitation is the finite number of scenarios considered in which disruption of a health system might occur. For example, we did not explored changes in sexual behaviour in the population due to disruption. However, we did account for a large variation in the duration of disruption and in vaccination coverage at disruption through our sensitivity analyses. In addition, we do not consider here the influence of choice of HPV vaccine, as a quadrivalent vaccine (either indigenously produced or Gardasil) would be the likely choice for India due to the price advantage. In cases of using a nonavalent vaccine, or a vaccine with higher levels of cross-protection, we expect similar qualitative results for resilience, and elimination of cervical cancer could be expected to be more easily achieved.

Although a formal health-economic assessment of the introduction of GN HPV vaccination in India is beyond the scope of this paper, clearly adding vaccination of boys to the routine coverage of girls would approximately double the required number of vaccine doses and would increase the financial effort to be made by the national government. However, it has been demonstrated that GN HPV vaccination schedules are economically attractive in high-income tender-based settings, in particular where GO vaccine uptake is below 80% (Qendri et al., 2020). GN vaccination is likely to be even more attractive in India where population-based screening programmes have not yet been widely implemented. Furthermore, the actual determinants of a trade-off between redundancy and efficiency of resource allocation depends on context-specific planning and assessment. In our example, to achieve greater resilience, GN vaccination with 60% coverage would require more doses of vaccine while leading to fewer prevented cases of cervical cancer in vaccinated cohorts without interruption than GO vaccination with 90% coverage (Figure 2). However, extending vaccination to boys could be easier to put in place than improving coverage in girls from 60% to 90% in some contexts. Finally, shifting from a two-dose to a single-dose HPV vaccination schedule could also improve the affordability of GN vaccination.

Having clearly illustrated the merits of GN vaccination considering different aspects across different settings here and in earlier studies (Chow et al., 2021; Elfstrom et al., 2016; Lehtinen et al., 2022; Lehtinen et al., 2018; Qendri et al., 2020), the next step is to establish a feasible pathway for implementation (de Sanjose & Bruni, 2020). In high-income countries, where affordability was not an issue, most vaccination programmes were initiated with the GO strategy, often combined with catch-up in girls/women, and are now increasingly shifting towards GN vaccination. In resource-limited settings, once routine vaccination in girls has been implemented, countries may need to make a choice between boys’ vaccination or female catch-up vaccination. That choice will depend on health-economic objectives, the type of health infrastructure already in place as well as societal values. In addition, shortages in vaccine supply will influence decision-making in the coming years until this is resolved (WHO, 2022b).

In conclusion, our model-based observations show that shifting from GO to GN vaccination may improve the resilience of the Indian HPV vaccination programme while also enhancing progress towards the elimination of cervical cancer. Our resilience estimates go beyond health-care disruption due to the COVID-19 pandemic. Indeed, any societal events undermining the routine activities of a healthcare system can produce a comparable effect. Over the years, disease prevention programmes have been disrupted by the occurrence of pandemics (Osterholm, 2017; WHO, 2021), armed conflicts (Jawad et al., 2021; McGrath, 2022), economic sanctions (Germani et al., 2022), or widespread vaccine hesitancy (Larson, 2020). Therefore, we argue that such societal crises, which are unpredictable but expected to occur, should be anticipated through careful planning of disease prevention programmes.

## Data Availability

External researchers can make written requests to the IARC for sharing of data regarding the IARC India HPV vaccine trial. Requests will be assessed on a case-by-case basis in consultation with lead investigators and co-investigators. A brief analysis plan and data request will be required and reviewed by the investigators for approval of data sharing. When requests are approved, anonymised data will be sent electronically
in password protected files. All data sharing will abide by rules and policies defined by the involved parties. Data sharing mechanisms will ensure that the rights and privacy of individuals participating in research will be protected at all times. Data are from public sources that are listed in the appendix (pp 6-14).

http://ci5.iarc.fr

https://www.ncdirindia.org/All_Reports/Report_2020/resources/NCRP_2020_2012_16.pdf

https://www.aidsdatahub.org/sites/default/files/resource/national-bss-general-population-india-2006.pdf

https://dhsprogram.com/

https://population.un.org/wpp/Download

https://censusindia.gov.in/2011census/C-series/C-13.html

## ACKNOWLEDGMENTS

This study was funded by the Bill & Melinda Gates Foundation (grant number: INV-039876). For the authors identified as personnel of the International Agency for Research on Cancer or World Health Organization, the authors alone are responsible for the views expressed in this article and they do not necessarily represent the decisions, policy or views of the International Agency for Research on Cancer or World Health Organization. The designations used and the presentation of the material in this Article do not imply the expression of any opinion whatsoever on the part of WHO and the IARC about the legal status of any country, territory, city, or area, or of its authorities, or concerning the delimitation of its frontiers or boundaries.

## DATA AVAILABILITY

Data for model calibration were publicly available from sources that are listed in the supplementary material of an earlier publication (pp 11–13, DOI: 10.1016/S1470-2045(22)00543-5). The model code of EpiMetHeos and Atlas are available from the authors on request.

## AUTHOR CONTRIBUTIONS

Supervision, Funding acquisition: IB, PB, SR

Conceptualization, Formal analysis, Writing – original draft: IB, IM

Methodology, Software, Visualization: IB, IM, DG

Validation, Writing – Review & Editing: IB, IM, DG, PB, SR

