## Appendices for "Building resilient cervical cancer prevention through gender-neutral HPV vaccination"

#### List of tables

Table A1. List of model parameters
Table A2. Overview of available cancer incidence data from local registries by Indian state
Table A3. Age-specific cervical cancer incidence data by Indian state
Table A4. Female mortality rate of India
Table A5. Type-specific contribution of HPV types in cervical cancer
Table A6. Standard world population
Table A7. Female population size by Indian state
Table A8. Pre-vaccination risk of cervical cancer by Indian state
Table A9. HPV-FRAME checklist

Table B1. Sensitivity analyses on coverage at disruption and duration of disruption on resilience by Indian state

#### List of figures

Figure A1. Structure of HPV natural history in EpiMetHeos
Figure A2. Model fit of the target statistics of sexual contact behaviour
Figure A3. Model fit of the type-specific HPV prevalence data
Figure A4. Age-specific cervical cancer incidence data by Indian state
Figure A5. Mean of age-specific cervical cancer incidence of the cluster of Indian states
Figure A6. Lexis diagram showing the 5-year birth cohorts targeted by HPV vaccination

Figure B1. Resilience against HPV vaccination disruption in the base case by Indian state
Figure B2. Resilience against HPV vaccination disruption in sensitivity analyses on coverage at disruption by Indian state
Figure B3. Resilience against HPV vaccination disruption in sensitivity analyses on duration of disruption by Indian state
Figure B4. Progress towards cervical cancer elimination over time with and without disruption

### Appendix A

#### A.1. HPV Transmission model

We adapted a previously published HPV transmission model[1] into an stochastic agent-based dynamic model, EpiMetHeos. EpiMetHeos is an extension of EpiModel[2] an open-source statistical framework that allows simulation of infectious disease transmission on dynamic contact networks.

##### A.1.1. Demography

The model considers an open population in which individuals enter at the age of 10 years and exit due to death with maximum age of 100 years. Each individual is characterised by the two demographic characteristics, age and sex, denoted by $a \in [10{,}100]$ and $g \in \{W, M\}$, respectively. The entire age range is further stratified into the following age groups: $ageg \in \{10-14, 15-19, 20-24, 25-29, 30-49, 50-59, 60-99\}$.

Individuals die according to sex- and age-specific probabilities $m_{g,age}$. The number of new individuals born into the population per time step is given by $b$, which is set to a value that keeps the total population constant over time and with 50%-50% of female and male new-borns. A constant age-specific population size distribution corresponding to $m_{g,age}$ is achieved through the simulation of a burn-in period.

##### A.1.2. Sexual contact behaviour

Two dynamic contact networks of sexual partnerships are modelled among the model individuals: one for stable partnerships and one for one-off partnerships. Stable partnerships represent marital partnerships and one-off partnerships outside marriage. The networks are constructed using EpiModel's implementation of the Separable Temporal Exponential-family Random Graph Models (STERGM).[2] An STERGM is characterised by formation and dissolution probabilities of partnerships. At each time step, partnerships that were present at the previous time step can be dissolved, and new partnership can be formed between individuals that were not connected at the previous time step.

Formation of partnerships is only allowed between opposite sex individuals to model heterosexual networks. Formation probabilities of stable as well as one-off partnerships depend on the individual's sex, age group and the assigned risk group of sexual activity. Each individual is assigned to one of the five risk groups of sexual activity, denoted by $riskg \in \{1,2,3,4,5\}$, which determines which type of partnerships he/she is allowed to form as follows:

- $riskg = 1$: no stable, no one-off,
- $riskg = 2$: only stable,
- $riskg = 3$: both stable and one-off,
- $riskg = 4$: only one-off,
- $riskg = 5$: only one-off; in women, this risk group represents female sex workers.

The risk group of an individual is assigned randomly at birth according to a sex-specific multinomial distribution $p_{g,riskg}$ and remains unchanged for the rest of the individual's lifespan. The dissolution probability differs between stable and one-off partnerships but is the same for all individuals.

Sexual behaviour is further characterised based on parameters regarding the number of sex acts within established partnerships. At each time step, sex acts may occur within a stable partnership. The number of sex acts is randomly generated according to a Poisson distribution with mean $r^{main}$. Within a one-off partnership, the number of sex acts $r^{one-off}$ is exactly one. The parameter values regarding sexual contact behaviour were fixed or obtained through a calibration step described in Section A.2.2.

##### A.1.3. HPV natural history

HR HPV types are assumed to be transmitted independently, governed by type-specific natural history parameters, including the probability of transmission, duration of infection and natural immunity. Transmission of all HR HPV types are assumed to follow the "Susceptible-Infected-Susceptible" dynamics in men and the "Susceptible-Infected-Removed/Immune-Susceptible" dynamics in women (**Figure A1**). Effects of HIV, smoking, and use of contraceptives, as risk factors of HPV transmission and cervical cancer progression, were not included in the model. We chose not to include them in the model. For HIV infection, this choice was based on the low HIV prevalence in India.[3] As for smoking and the use of contraceptives, the choice was based on the lack of data.

All individuals enter the population being susceptible. At each time step, an unvaccinated susceptible individual having $l$ sex acts with an individual infected with type $i$ has a probability of $1-(1-\beta_i)^l$ becoming infected, where $\beta_i$ is the probability of transmission per sex act. If vaccinated and vaccine protection is successfully induced, the probability of transmission for a vaccine-targeted type, either a vaccine type or a cross-protective type, becomes zero. In other words, we assumed an all-or-nothing working mechanism for vaccine protection. When vaccine-induced protection is lost, the probability of transmission returns to $1-(1-\beta_i)^l$.

The duration of infection follows a type-specific distribution with six parameters $\gamma_i, \eta_i, \delta_{i,1}, \delta_{i,2}, \nu_{i,1}, \nu_{i,2}$. This distribution describes the process through CIN0, CIN1, regressive CIN2/3, and non-regressive CIN2/3 stages as modelled in an extensively validated cervical cancer progression model.[4,5] In particular, non-regressive CIN2/3 here represents the part of HPV infections that will persist and progress to cancer. For those HPV infections that do not persist, upon clearance, men become susceptible and women become removed/immune. The duration of natural immunity in women follows an exponential distribution type-specific rate $\mu_i$, after which women also become susceptible again. The values of the natural history parameters were fixed or obtained through a calibration step described in Section A.2.3 and are reported in **Table A1**.

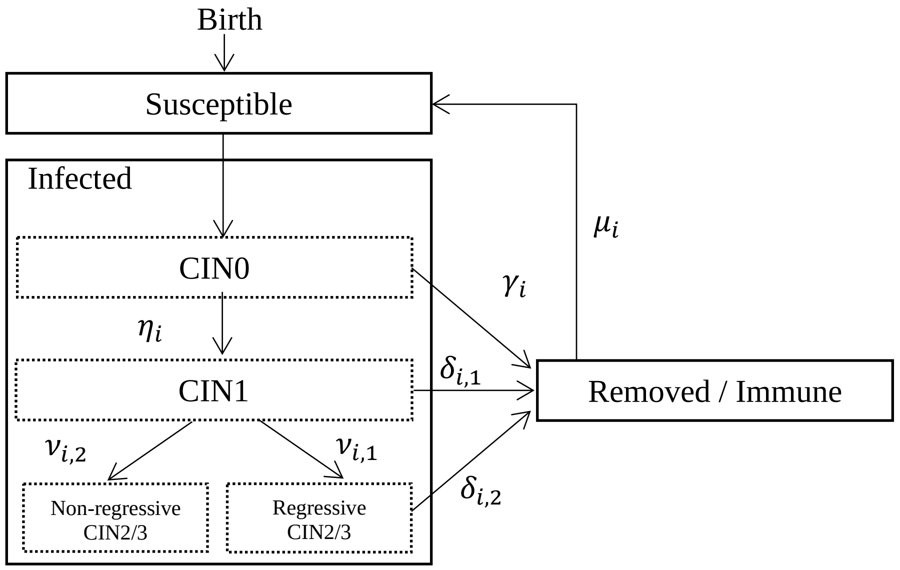

**Figure A1. Structure of HPV natural history in EpiMetHeos.**

#### A.2. Model calibration

##### A.2.1. Footprinting framework

Since high-quality data about HPV prevalence and cervical cancer incidence data essential to project the impact of HPV vaccination are not available for each Indian state, the Footprinting framework was used to approximate missing data and extrapolate impact projections. The Footprinting framework is described in more details in a separate manuscript.[6] The framework consists of three steps: clustering, classification, and projection. These steps are briefly described below.

Clustering step

In the clustering step, Indian states with available cervical cancer incidence data were clustered based on their similarity in patterns of age-specific cervical cancer incidence. Sources for cervical cancer incidence data were volume XI of Cancer Incidence in Five Continent (CI5) and the 2012-2016 Report of National Cancer Registry Programme by National Centre for Disease Informatics and Research (NCDIR).[7,8] See **Table A2** for an overview of Indian states (or groups of states) with available cancer incidence data from local registries. Of the 25 Indian states, 14 states have local registries and 11 do not. Whenever a registry was both present in CI5 and NCDIR, only the data by CI5 were used. See **Table A3** and **Figure A5** for the extracted age-specific cervical cancer incidence data.

A Poisson-regression-based CEM clustering algorithm was used to cluster the age-specific cervical cancer incidence curve. Details of the clustering algorithm are described in Appendix S1 of the separate manuscript are reported in the main text of the separate manuscript.[6] We identified one cluster with high cancer incidence and one with low cancer incidence. See also column “cluster” of **Table A3** for the assigned cluster of each Indian state. In this table, the Indian states with cervical cancer incidence, hence involved in this clustering step, are indicated by “extracted” in the column “source”.

Classification step

The remaining Indian states without cervical cancer incidence data were classified to the identified clusters of Indian states with similar patterns of cervical cancer incidence based on similarity in sexual behaviour data from the behaviour surveillance survey by the National AIDS Control Organization of India.[9]

Random forest was used for classification. Details of the classification method are described in Appendix S2 of the separate manuscript are reported in the main text of the separate manuscript.[6] See also column “cluster” of **Table A3** for the assigned cluster of each Indian state. In this table, the Indian states without cervical cancer incidence, hence involved in this classification step, are indicated by “inferred” in the column “source”.

Projection step

In the projection step, baseline (i.e., in the scenario without vaccination) HPV prevalence and cervical cancer incidence were approximated, based on the available data within each cluster. For HPV prevalence, high-quality type- and age-specific HPV prevalence data were only available for Tamil Nadu and West Bengal, the former being in the “high” and the latter in the “low” incidence cluster.[10,11] As for cervical cancer incidence data, approximation was based on the mean age-specific incidence within each cluster. See **Figure A5** and **Table A3** for mean values. From the approximated Indian state-specific baseline cervical cancer incidence, we then derived cervical cancer risk in terms of three indicators, lifetime risk (LTR), and age-standardized incidence rate (ASIR), according to the methodology described in **Sections A.3.2**, and **A.3.3**, respectively. See **Table A8** for baseline cervical cancer risk by LTR and ASIR.

HPV incidence and cervical cancer risk under vaccination scenarios were obtained as follows. The HPV transmission model, EpiMetHeos, was calibrated to these two representative Indian states (Tamil Nadu and West Bengal) according to the procedure described in **Section A.2.2.** Using the two obtained models, we then simulated different vaccination scenarios and obtained estimates of relative reduction in HPV infection risk according to the methodology described in **Sections A.3.1**. Finally, these estimates of relative reduction were applied to the previously obtained baseline values of cervical cancer risk to derive the values under vaccination scenarios.

##### A.2.2. Calibration to sexual behaviour data

The model was calibrated to the two representation states: West Bengal and Tamil Nadu. Model calibration consists of two steps. In the first step, the component of the model simulating the dynamic contact networks was calibrated to the sexual behaviour data without considering the component concerning the natural history of HPV yet. Four sources of sexual behaviour data were used:

- The Demographic and Health Survey (DHS) programme, to provide information on stable partnerships in West Bengal and Tamil Nadu.[12]
- National Behavioural Surveillance Survey 2006 by The National AIDS Control Organisation (NACO), to provide information on one-off partnerships in West Bengal and Tamil Nadu.[9]
- Publication by Gaffey et al., “Male use of female sex work in India: a nationally representative behavioural survey”, to provide information on proportion of men with stable partnership that also have one-off partnerships in West Bengal and Tamil Nadu.[13]
- Publication by Vandepitte et al, “Estimates of the number of female sex workers in different regions of the world”, to provide information on one-off partnerships across India.[14]

First, the probabilities of being assigned to different risk groups of sexual activity were fixed. See **Table A1** for the fixed values and the justification. Subsequently, formation and dissolution probabilities of the dynamic sexual contact networks were fitted to a set of target statistics using the *netest* function of EpiModel. The set of target statistics was:

- Stable partnerships:
  - Sex- and age-group-specific population proportion with stable partnerships $d_{g,ageg}^{stable}$
  - Sex- and risk-group-specific population proportion with stable partnerships $d_{g,riskg}^{stable}$
  - Overall population proportion with stable partnerships $d^{stable}$
  - Mean age difference in stable partnerships $\kappa^{stable}$
  - Mean spread of absolute age difference in stable partnerships $\omega^{main}$
  - Mean duration of stable partnerships $s^{stable}$
- One-off partnerships:
  - Sex- and age-group-specific mean degree of one-off partnerships $d_{g,ageg}^{one-off}$
  - Sex- and risk-group-specific mean degree of one-off partnerships $d_{g,riskg}^{one-off}$
  - Overall mean degree of one-off partnerships $d^{one-off}$
  - Mean duration of one-off partnerships $s^{one-off}$

See **Table A1** for the values of the target statistics and the corresponding data sources used. See **Table A1** also for the fixed parameters and the corresponding data sources used. See **Figure A2** for the fit to the target statistics. Note that assortativeness of sexual contact by age- and risk-groups are results of the fitting process.

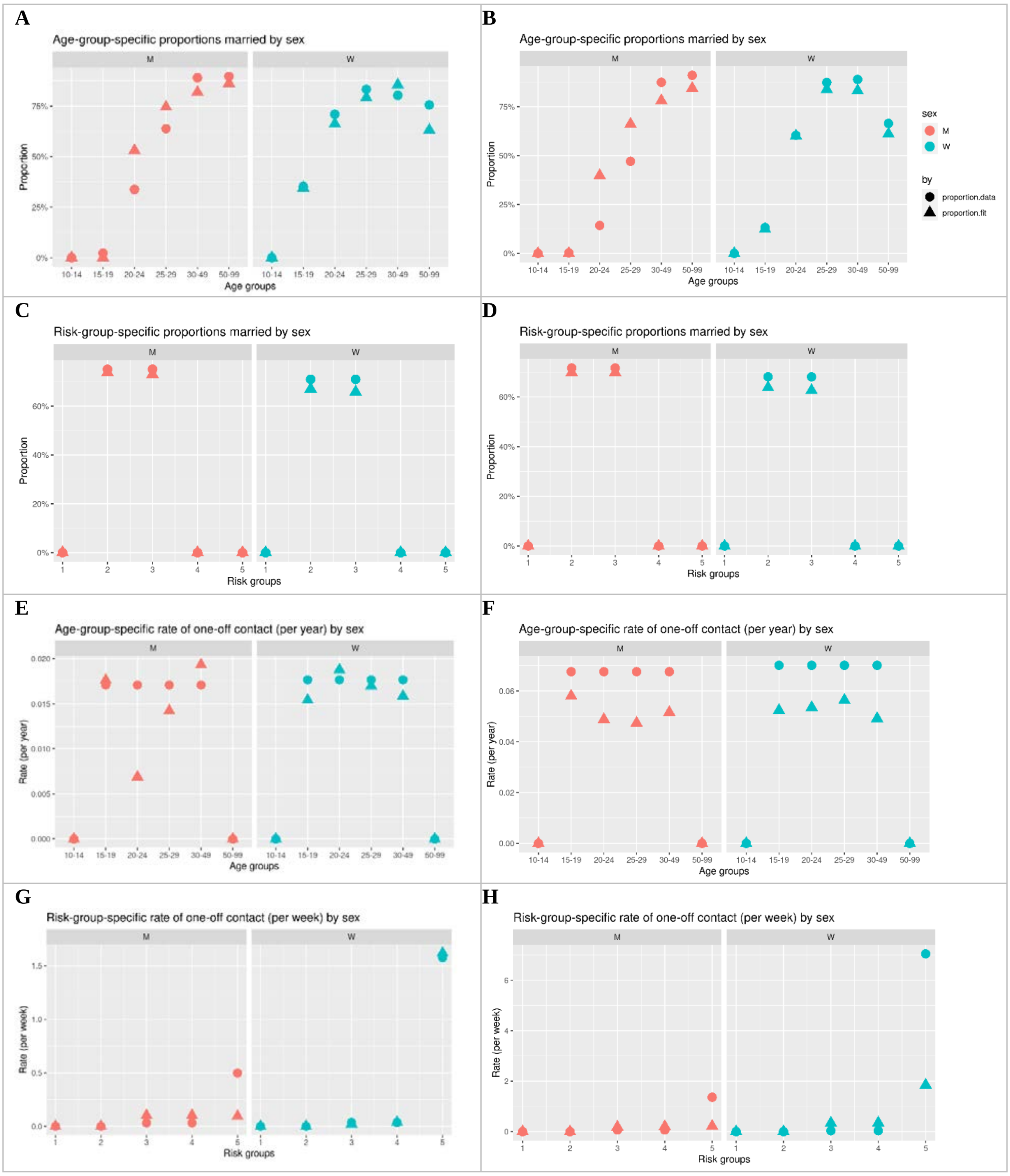

**Figure A2. Model fit of the target statistics of sexual contact behaviour**. Left column: West Bengal. Right column: Tamil Nadu. Circle: targets derived from data. Triangle: fit by model.

**A.2.3. Calibrating to HPV prevalence data**

In the second step, the entire model was calibrated, including the component concerning the natural history of HPV. This was done by fitting the model HPV prevalence among sexually active women to HPV prevalence data from two surveys among married women in Tamil Nadu and West Bengal.[10,11] To obtain consistent type-specific HPV natural history parameters between Tamil Nadu and West Bengal, we scaled the relative type-specific prevalence of Tamil Nadu to West Bengal. The relative prevalence of HPV 16/18 between Tamil Nadu and West Bengal was used as the scaling factor.

Due to the low prevalence of some HR HPV types, we used the average prevalence of the following three groups of HR HPV types as target prevalence:

- HPV 16
- HPV 18
- HPV 31/33/45/35/39/51/52/56/58/59/68

Note that for the subsequent simulation of the vaccination scenarios, the calibrated parameters based on average prevalence of HPV 31/33/45/35/39/51/52/56/58/59/68 were then used to model the cross-protective types HPV 31/33/45 and the other HR HPV types HPV 35/39/51/52/56/58/59/68, which are referred to as "*cross*" and "*other*".

The parameter values regarding the type-specific progression, clearance rates and waning rates of natural immunity were fixed to those estimated for the extensively validated cervical cancer progression model.[4,5] For HPV 31/33/45/35/39/51/52/56/58/59/68, we derived the mean values of the type-specific estimates.

The parameter values obtained in this calibration step regard type-specific transmission probabilities and one-off partnership underreporting rate. Using 2500 parameter sets that were uniformly generated from the range (**Table A1**), we selected 100 best-fitting parameter sets. Model fit was evaluated based on log-likelihood of the observed HPV prevalence data given the simulated HPV prevalence under a binomial distribution. For each parameter set, log-likelihood was computed at each year in the last 50 years of 250 years of simulation. The maximum and mean log-likelihood across these years were used as the two summary statistics. See **Figure A3** for the fit to the HPV target prevalence.

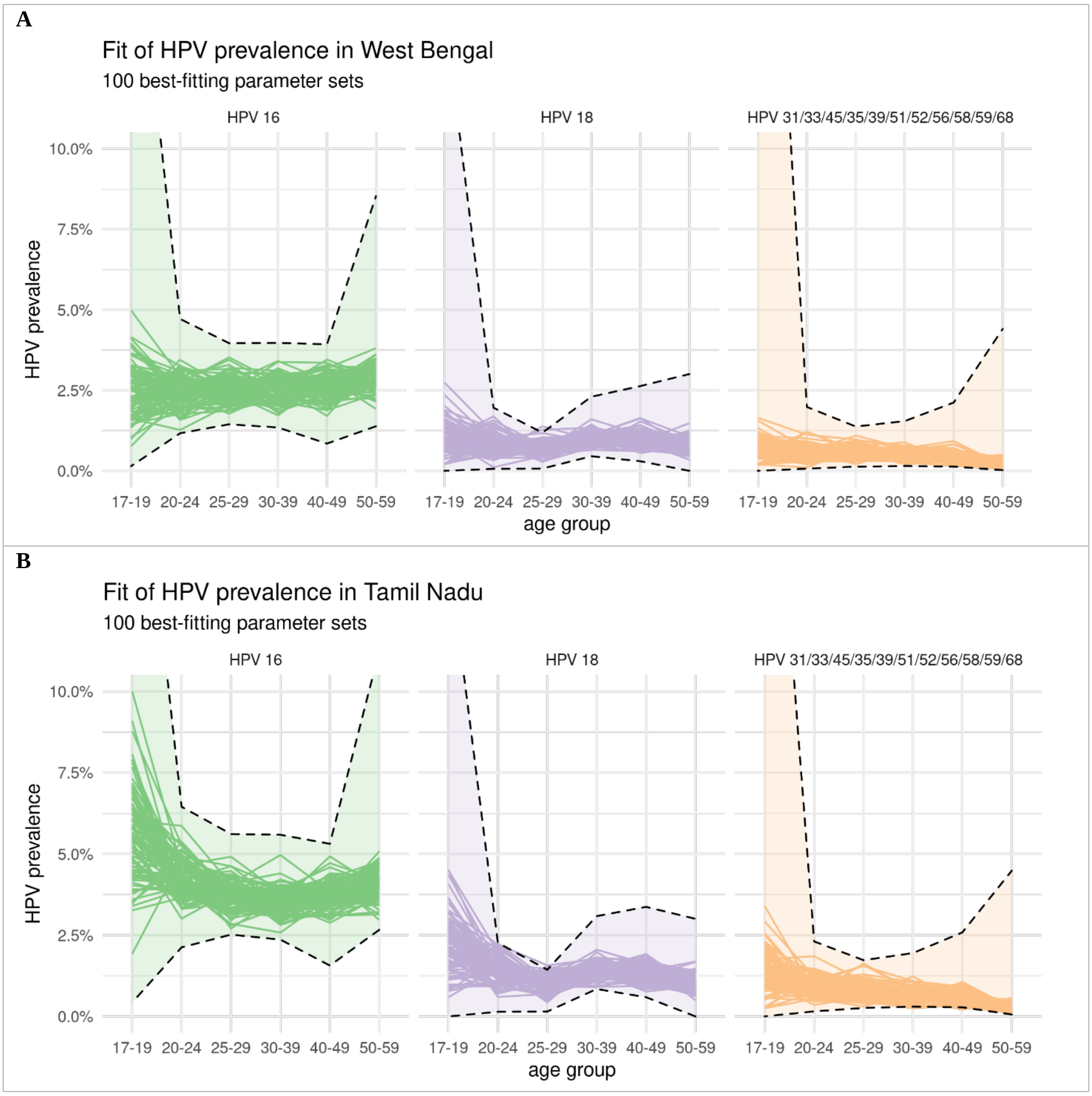

**Figure A3. Model fit of the type-specific HPV prevalence data**. Panel A: West Bengal. Panel B: Tamil Nadu. Model estimates of HPV prevalence of each of the 100 best-fitting parameter sets are given by a separate line. The confidence intervals of the observed HPV prevalence under binomial distribution are given by the dashed lines. The confidence intervals for West Bengal follow the same shape as those for Tamil Nadu, as the type-specific prevalence for West Bengal was derived by rescaling that of Tamil Nadu (the prevalence for West Bengal being approximately half that of Tamil Nadu).

**Table A1. List of model parameters.**

| Notation | Description | Values / ranges | Reference |
|---|---|---|---|
| *Demography* | | | |
| $m_{g,age}$ | Gender- and age-specific mortality | See **Table A4** | Based UN life tables of India in 2015-2020.[15] |
| $b$ | Population sex-specific birth rate | Set such that the total population stays constant given $m_{g,age}$ and 50%-50% distribution of female and male new-borns | NA |
| *Sexual contact behaviour* | | | |
| $p_{W,1}, p_{M,1}$ | Probability of being assigned to risk group 1 in women and men | 1.3%, 3.3% for West Bengal<br>1.7%, 2.8% for Tamil Nadu | Proportion of virgins in the age group 30-49 estimated by DHS in West Bengal and Tamil Nadu.[12] |
| $p_{M,3}, p_{M,4}, p_{M,5}$ | Probability of being assigned to risk group 3, 4, and 5 in men | 3.6%, 4.7%, 1.4% for West Bengal<br>7.9%, 4.6%, 2.2% for Tamil Nadu | Based on Table 1 of Gaffey et al.[13] The rows for low- and high-HIV states were used for West Bengal and Tamil Nadu, respectively. Risk groups 3, 4 and 5 were based on the proportion of sexually active married men having non-regular partners, sexually active unmarried men having non-regular partners but not using female sex workers, and sexually active unmarried men using female sex worker, respectively. |
| $p_{W,5}$ | Probability of being assigned to risk group 5 in women, representing female sex workers | 0.5% for West Bengal and Tamil Nadu | Estimated prevalence of female sex workers for India in Table 2 of Vandepitte et al.[14] |
| $p_{W,3}, p_{W,4}$ | Probability of being assigned to risk groups 3 and 4 in women | 2.5%, 2.5% for West Bengal<br>2.65%, 2.65% for Tamil Nadu | Urban percentage of women with at least one non-regular partners last year after subtracting the proportion of female sex workers in Table 6.2 of the NACO report.[9] |
| $p_{W,2}, p_{M,2}$ | Probability of being assigned to risk group 2 in women and men | 86.0%, 93.2% for West Bengal<br>82.5%, 92.5% for Tamil Nadu | Remaining proportion after fixing the proportions of other risk groups. |
| $d^{stable}_{W,10-14}$, $d^{stable}_{M,10-14}$ | Population proportion with stable partnerships in women and men aged in the age group 10-14 | 0% | Set as zero given the low proportion of married individuals in this age groups as estimated by DHS.[12] |
| $d^{stable}_{W,15-19}, d^{stable}_{W,20-24}, d^{stable}_{W,25-29}, d^{stable}_{W,30-49}, d^{stable}_{W,50-59}, d^{stable}_{W,60-99}$ | Population proportion with stable partnerships in women in age groups 15-19, 20-24, 25-29, 30-49, 50-59, and 60-99 | 38.5%, 77.6%, 91.1%, 87.8%, 83%, 83% for West Bengal<br>12.8%, 58.5%, 84.7%, 86.2%, 64%, 64% for Tamil Nadu | Proportion of women married in DHS in West Bengal and Tamil Nadu.[12] |
| $d^{stable}_{M,15-19}, d^{stable}_{M,20-24}, d^{stable}_{M,25-29}, d^{stable}_{M,30-49}, d^{stable}_{M,50-59}, d^{stable}_{M,60-99}$ | Age-group-specific population proportion with stable partnerships in men in age groups 15-19, 20-24, 25-29, 30-49, 50-59, and 60-99 | 2.4%, 34.8%, 65.8%, 91.6%, 92.3, 92.3% for West Bengal<br>0.3%, 15.2%, 50.1%, 93.1%, 96.9, 96.9% for Tamil Nadu | Proportion of women married in DHS in West Bengal and Tamil Nadu.[12] |
| $d^{stable}_{W,1}, d^{stable}_{W,4}, d^{stable}_{W,5}, d^{stable}_{M,1}, d^{stable}_{M,4}, d^{stable}_{M,5}$ | Population proportion with stable partnerships in risk groups 1, 4, and 5 | 0%, by definition. | NA |
| $d^{stable}_{W,2}, d^{stable}_{W,3}$ $d^{stable}_{M,2}, d^{stable}_{M,3}$ | Population proportion with stable partnerships in risk groups 2 and 3 | Set to match the age-group-specific population proportion with stable partnerships:<br>75% for West Bengal | NA |

| | | | |
|---|---|---|---|
| | | 72% for Tamil Nadu | |
| $d^{stable}$ | Overall population proportion with stable partnerships | Set to match the age-group-specific population proportion with stable partnerships:<br>68% for West Bengal<br>65% for Tamil Nadu | NA |
| $\kappa^{stable}$ | Mean age difference in stable partnerships (men minus women) | 7.29 years for West Bengal<br>6.58 years for Tamil Nadu | Estimated from DHS for West Bengal and Tamil Nadu.[12] |
| $\omega^{main}$ | Mean spread of absolute age difference in stable partnerships | 3.31 years for West Bengal<br>3.28 years for Tamil Nadu | Estimated from DHS for West Bengal and Tamil Nadu.[12] |
| $s^{stable}$ | Mean duration of stable partnerships | 20 years, by assumption. | NA |
| $d_{W,1}^{one-off}, d_{W,2}^{one-off}, d_{M,1}^{one-off}, d_{M,2}^{one-off}$ | Mean degree of one-off partnerships in risk groups 1 and 2 | 0%, by definition. | NA |
| $d_{M,3}^{one-off}, d_{M,4}^{one-off}, d_{M,5}^{one-off}$ | Mean degree of one-off partnerships in risk groups 3, 4, and 5 in men | 1.65, 1.65, 25.57 for West Bengal<br>3.41, 3.41, 70.58 for Tamil Nadu | Derived on in Table 6.4 of the NACO report.[9] |
| $d_{W,3}^{one-off}, d_{W,4}^{one-off}$ | Mean degree of one-off partnerships in risk groups 3 and 4 in women | 2, by assumption. | NA |
| $d_{W,5}^{one-off}$ | Sex- and age-group-specific mean degree of one-off partnerships | Set to match the degree of one-off partnerships in men:<br>89.1 partners per year in West Bengal<br>373.4 partners per year in Tamil Nadu | NA |
| $d^{one-off}$ | Overall mean degree of one-off partnerships | Set to match the risk-group-specific degree with one-off partnerships:<br>0.50 partners per year for West Bengal<br>1.98 partners per year for Tamil Nadu | NA |
| $s^{one-off}$ | Mean duration of one-off partnerships | 1 unit of time step, by definition | NA |
| $r^{stable}$ | Mean number of sex acts per time step within stable partnerships according to a Poisson distribution | | |
| $r^{one-off}$ | Fixed number of sex acts per time step within one-off partnerships | 1 act, by definition | NA |
| $\theta^{one-off}$ | One-off partnership underreporting rate | Derived from model calibration with the following range for candidate values: (8, 20) | NA |
| *HPV natural history* | | | |
| $\beta_i$ | Transmission probability of type $i$ per sex act | Derived from model calibration with the following range for candidate values: (0.2, 0.95). | NA |
| $\gamma_{16}, \gamma_{18}, \gamma_{cross}, \gamma_{other}$ | Clearance rate from CIN0 | 0.824 per year for HPV 16,<br>0.955 per year for HPV 18,<br>1.18 per year for the cross-protective types and remaining HR HPV types | [4,5] |
| $\eta_{16}, \eta_{18}, \eta_{cross}, \eta_{other}$ | Progression rate from CIN0 to CIN1 | 0.676 per year for HPV 16,<br>0.545 per year for HPV 18, | [4,5] |

| | | | |
|---|---|---|---|
| | | 0.324 per year for the cross-protective types and remaining HR HPV types | |
| $\delta_{16,1}, \delta_{18,1}, \delta_{cross,1}, \delta_{other,1}$ | Clearance rate from CIN1 | 0.133 per year for HPV 16,<br>0.386 per year for HPV 18,<br>0.481 per year for the cross-protective types and remaining HR HPV types | [4,5] |
| $\delta_{16,2}, \delta_{18,2}, \delta_{cross,2}, \delta_{cross,2}$ | Clearance rate from regressive CIN2/3 | 2.10 per year for HPV 16,<br>2.10 per year for HPV 18,<br>2.10 per year for the cross-protective types and remaining HR HPV types | [4,5] |
| $\nu_{16,1}, \nu_{18,1}, \nu_{cross,1}, \nu_{other,1}$ | Progression rate from CIN1 to regressive CIN2/3 | 0.048 per year for HPV 16,<br>0.00681 per year for HPV 18,<br>0.0447 per year for the cross-protective types and remaining HR HPV types | [4,5] |
| $\nu_{16,2}, \nu_{18,2}, \nu_{cross,2}, \nu_{other,2}$ | Progression rate from CIN1 to non-regressive CIN2/3 | 0.0454 per year for HPV 16,<br>0.0450 per year for HPV 18,<br>0.0110 per year for the cross-protective types and remaining HR HPV types | [4,5] |
| $\mu_{16}, \mu_{18}, \mu_{cross}, \mu_{other}$ | Rate of waning natural immunity | 0.0407 per year for HPV 16,<br>0.0287 per year for HPV 18,<br>0.0320 per year for the cross-protective types and remaining HR HPV types | [4,5] |

#### A.3. Computation of model outcomes

##### A.3.1. Cumulative risk of HPV infection

For each of the four representative HPV type $i$ and each birth cohort, model estimates on half-year incidence $r_{i,a}$ up to age $a = 40$ years were combined to derive the cumulative risk of HPV infection $Y_i$, as follows: $Y_i = \sum_{a=15}^{45} 0.5 r_{i,a} s_a$. Here, $s_a = \exp\,(-\sum_{a'=10}^{4} 0.5 d_{a'})$ is the survival probability up to age $a$ derived from the mortality rates $d_{a'}$ of the UN life-table of India in 2015-2020 (**Table A4**).

The cumulative risk of any HR HPV infection was a weighted average of the relative reduction in type-specific cumulative risk of HPV infection. The weights were based on the type-specific contribution to cervical cancer as observed in India.[16] The contributions by HPV 16, 18, and 31/33/45 were 62%, 16%, and 6%, respectively (**Table A5**). These proportions were derived by normalizing the type-specific contributions.

As for outcome of HPV prevalence, we assumed the model estimates for cumulative risk of HPV infection for West Bengal and Tamil Nadu to apply to all states in the low- and high-cancer-incidence clusters, respectively.

##### A.3.2. Life-time risk of cervical cancer

To obtain the life-time risk (LTR) of cervical cancer in the scenario without vaccination, we extracted age-specific cervical cancer incidence data from volume XI of Cancer Incidence in Five Continent (CI5) and the 2012-2016 Report of National Cancer Registry Programme by National Centre for Disease Informatics and Research (NCDIR).[7,8] Of the 25 Indian states, 14 states have local registries and 11 do not. Whenever a registry was present in both CI5 and NCDIR, only the data corresponding to CI5 were included. As described in **Section A.2.1**, each state without a local cancer registry was classified to either the cluster of states with high or low cervical cancer incidence. The missing incidence were inferred based on the cluster mean of the classified cluster. See **Table A3**, **Figure A4** and **Figure A5** for the extracted or inferred age-specific cervical cancer incidence data by Indian state.

A recently published method[17] was used to derive LTR of cervical cancer from age-specific incidence. This method converts age-specific cancer incidence into LTR of cervical cancer (in cases per 100,000 girls born) while accounting for the competing risk of dying from other causes before possible occurrence of cervical cancer. For LTR, we used the UN data on female mortality rates for 2015-2020 in India (**Table A4**).[15] See **Table A8** for the baseline LTR without vaccination by Indian state.

To derive the LTR of cervical cancer in the scenarios with vaccination, we approximated the relative reduction in risk of cervical cancer by model estimates of the relative reduction in the cumulative risk of any HR HPV infection. Model estimates of West Bengal and Tamil Nadu were used for the states in the low- and high-cancer-incidence clusters, respectively.

##### A.3.3. Age-standardised incidence rate of cervical cancer

Age-standardised incidence rate (ASIR) of cervical cancer (in cases per 100,000 woman-years) was obtained based on the world standard population (**Table A6**).[18] See **Table A8** for the baseline ASIR without vaccination by state. As for the LTR, the ASIR in the scenarios with vaccination was derived based on model estimates of the relative reduction in the cumulative risk of any HR HPV infection.

##### A.3.4. Aggregating model outcomes

To obtain outcomes for India as a whole and by low- and high-incidence states, state-specific outcomes were weighted based on the state-specific female population size according to the table C-13 by the Indian Census of 2011 (**Table A7**).[19]

Model outcomes corresponding to the simulation of the 100 best-fitting parameter sets were used to derive the mean and the 10th and 90th percentiles, i.e., Uncertainty Interval (UI), of the model outcomes. For the nationwide model outcomes, the 100 best-fitting parameter sets of West Bengal and Tamil Nadu were paired for derived 100x100 outcomes, which were subsequently used to derive the mean and the 10th and 90th percentiles. Furthermore, nationwide outcomes and outcomes across all high- and low-cancer-incidence states were obtained by weighting the state-specific outcomes by the corresponding population sizes.

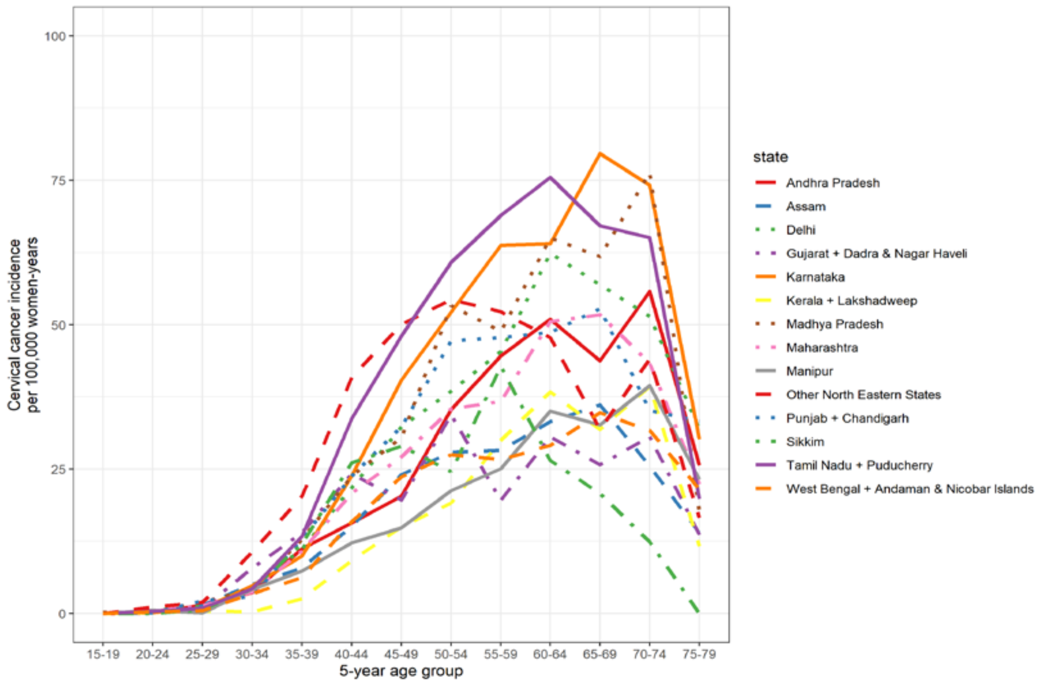

**Figure A4. Age-specific cervical cancer incidence data by Indian state.**

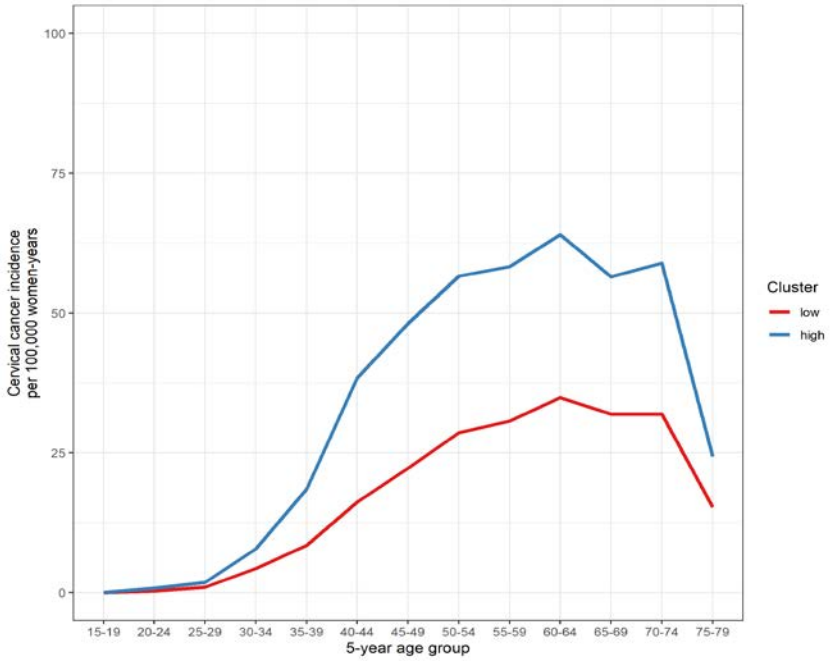

**Figure A5. Mean of age-specific cervical cancer incidence of the cluster of Indian states**

**Table A2. Overview of available cancer incidence data from local registries by Indian state.**
* States or groups of states as reported in the 2006 National Behaviour Surveillance Survey of the National AIDS Control Organization of India.[9]
§ Other North Eastern States include Arunachal Pradesh, Nagaland, Meghalaya, Mizoram, and Tripura.
¤ The eighteen registries CI5 and NCDIR do not have in common are in *italics*.

| State/group of states * | CI5 registry ¤ [7] | NCDIR registry ¤ [8] |
|---|---|---|
| Andhra Pradesh | | *Hyderabad district* |
| Assam | Cachar, Kamrup Urban District | Cachar district, *Dibrugarh district*, Kamrup urban |
| Bihar | | |
| Chhattisgarh | | |
| Delhi | | *Delhi* |
| Goa + Daman & Diu | | |
| Gujarat + Dadra & Nagar Haveli | Ahmedabad | Ahmedabad urban |
| Haryana | | |
| Himachal Pradesh | | |
| Jammu & Kashmir | | |
| Jharkhand | | |
| Karnataka | Bangalore | Bangalore |
| Kerala + Lakshadweep | Kollam, Trivandrum | Kollam district, Thi'puram district |
| Madhya Pradesh | Bhopal | Bhopal |
| Maharashtra | Barshi & Paranda & Bhum, Mumbai, Poona, Wardha | *Aurangabad*, *Osamanabad & Beed*, Barshi rural, Mumbai, Pune, Wardha district, *Nagpur* |
| Manipur | | *Manipur state*, *Imphal West district* |
| Orissa | | |
| Other North Eastern States § | Mizoram, Tripura | Mizoram state, *Aizawl district*, Tripura state, *West Arunachal*, *Papumpare district*, *Meghalaya*, *East Khasi Hills district*, *Nagaland*, *Pasighat* |
| Punjab + Chandigarh | | *Patiala district* |
| Rajasthan | | |
| Sikkim | Sikkim State | Sikkim state |
| Tamil Nadu + Puducherry | Chennai, *Dindigul Ambilikkai* | Chennai |
| Uttar Pradesh | | |
| Uttarakhand | | |
| West Bengal + Andaman & Nicobar Islands | | *Kolkata* |

**Table A3. Age-specific cervical cancer incidence data by Indian state.**
Incidence is given in cases per 100,000 woman-years by 5-year age groups.
* States or groups of states as reported in the 2006 National Behaviour Surveillance Survey of the National AIDS Control Organization of India.[9]
§ Other North Eastern States include Arunachal Pradesh, Nagaland, Meghalaya, Mizoram, and Tripura.
₮ "Extracted": cervical cancer incidence data were extracted from CI5 or NCDIR when available; "Inferred": when cervical cancer incidence data were unavailable, they were inferred based on footprinting.[6-8]
¤ Belonging to low- or high-incidence cluster. Cluster was obtained by the clustering step when cervical cancer incidence data were available and by the classification step whenever cervical cancer incidence data were unavailable.

| | | | Age group | | | | | | | | | | | | | |
|---|---|---|---|---|---|---|---|---|---|---|---|---|---|---|---|---|
| **State/group of states *** | **Source ₮** | **Cluster ¤** | **15-19** | **20-24** | **25-29** | **30-34** | **35-39** | **40-44** | **45-49** | **50-54** | **55-59** | **60-64** | **65-69** | **70-74** | **75-79** | **80-84** |
| Andhra Pradesh | Extracted | Low | 0 | 0.1 | 1.5 | 3.6 | 11.2 | 15.7 | 20.3 | 35.3 | 44.6 | 51 | 43.8 | 55.7 | 25.7 | 12.8 |
| Assam | Extracted | Low | 0 | 0.2 | 1.5 | 5.1 | 7.9 | 15.1 | 24.1 | 27.9 | 28.2 | 33.2 | 36.1 | 25.5 | 13.9 | 7 |
| Bihar | Inferred | Low | 0 | 0.3 | 1 | 4.3 | 8.4 | 16.2 | 22.2 | 28.6 | 30.7 | 34.9 | 31.9 | 31.9 | 15.3 | 7.6 |
| Chhattisgarh | Inferred | Low | 0 | 0.3 | 1 | 4.3 | 8.4 | 16.2 | 22.2 | 28.6 | 30.7 | 34.9 | 31.9 | 31.9 | 15.3 | 7.6 |
| Delhi | Extracted | High | 0 | 0.6 | 1.2 | 4.4 | 11.2 | 21.8 | 32.3 | 38.5 | 45.4 | 62.3 | 57 | 51.4 | 31.9 | 16 |
| Goa + Daman & Diu | Inferred | Low | 0 | 0.3 | 1 | 4.3 | 8.4 | 16.2 | 22.2 | 28.6 | 30.7 | 34.9 | 31.9 | 31.9 | 15.3 | 7.6 |
| Gujarat + Dadra & Nagar Haveli | Extracted | Low | 0 | 0 | 0.8 | 7.9 | 14 | 24.2 | 19.7 | 34.4 | 19.6 | 30.6 | 25.7 | 30.6 | 13.6 | 6.8 |
| Haryana | Inferred | Low | 0 | 0.3 | 1 | 4.3 | 8.4 | 16.2 | 22.2 | 28.6 | 30.7 | 34.9 | 31.9 | 31.9 | 15.3 | 7.6 |
| Himachal Pradesh | Inferred | Low | 0 | 0.3 | 1 | 4.3 | 8.4 | 16.2 | 22.2 | 28.6 | 30.7 | 34.9 | 31.9 | 31.9 | 15.3 | 7.6 |
| Jammu & Kashmir | Inferred | Low | 0 | 0.3 | 1 | 4.3 | 8.4 | 16.2 | 22.2 | 28.6 | 30.7 | 34.9 | 31.9 | 31.9 | 15.3 | 7.6 |
| Jharkhand | Inferred | Low | 0 | 0.3 | 1 | 4.3 | 8.4 | 16.2 | 22.2 | 28.6 | 30.7 | 34.9 | 31.9 | 31.9 | 15.3 | 7.6 |
| Karnataka | Extracted | High | 0 | 0.2 | 0.9 | 4.9 | 10 | 23.8 | 40.4 | 52.2 | 63.7 | 64 | 79.6 | 74.1 | 30.2 | 15.1 |
| Kerala + Lakshadweep | Extracted | Low | 0 | 0.2 | 0.4 | 0.3 | 2.5 | 9.2 | 15 | 19.1 | 30 | 38.4 | 31.9 | 39.3 | 11.7 | 5.8 |
| Madhya Pradesh | Extracted | High | 0.2 | 0.2 | 2 | 4.3 | 12.8 | 23.8 | 30.4 | 53.3 | 48.8 | 65 | 61.8 | 76.4 | 17.7 | 8.8 |
| Maharashtra | Extracted | Low | 0 | 0.4 | 1.3 | 3.8 | 10.8 | 20.8 | 27.1 | 35.4 | 36.8 | 50.5 | 51.7 | 43.3 | 22.4 | 11.2 |
| Manipur | Extracted | Low | 0 | 0.6 | 0.1 | 4.2 | 7.4 | 12.2 | 14.8 | 21.3 | 25.1 | 35 | 32.7 | 39.4 | 23.2 | 11.6 |
| Orissa | Inferred | Low | 0 | 0.3 | 1 | 4.3 | 8.4 | 16.2 | 22.2 | 28.6 | 30.7 | 34.9 | 31.9 | 31.9 | 15.3 | 7.6 |
| Other North Eastern States § | Extracted | High | 0 | 1.1 | 1.9 | 10.7 | 20.3 | 40.9 | 50 | 54.3 | 52.2 | 47.8 | 31.9 | 44 | 16.6 | 8.3 |
| Punjab + Chandigarh | Extracted | High | 0 | 0.1 | 2.2 | 3 | 13 | 23.8 | 32.3 | 47.2 | 47.8 | 48.7 | 52.8 | 35.2 | 33.3 | 16.6 |
| Rajasthan | Inferred | Low | 0 | 0.3 | 1 | 4.3 | 8.4 | 16.2 | 22.2 | 28.6 | 30.7 | 34.9 | 31.9 | 31.9 | 15.3 | 7.6 |
| Sikkim | Extracted | Low | 0 | 0 | 0.7 | 4.3 | 12 | 26.1 | 29 | 24.6 | 42.8 | 26.5 | 20.8 | 12.4 | 0 | 0 |
| Tamil Nadu + Puducherry | Extracted | High | 0 | 0.4 | 0.9 | 4.3 | 13.4 | 33.7 | 48.1 | 60.8 | 68.9 | 75.5 | 67.1 | 65.1 | 19.9 | 9.9 |
| Uttar Pradesh | Inferred | Low | 0 | 0.3 | 1 | 4.3 | 8.4 | 16.2 | 22.2 | 28.6 | 30.7 | 34.9 | 31.9 | 31.9 | 15.3 | 7.6 |
| Uttarakhand | Inferred | Low | 0 | 0.3 | 1 | 4.3 | 8.4 | 16.2 | 22.2 | 28.6 | 30.7 | 34.9 | 31.9 | 31.9 | 15.3 | 7.6 |
| West Bengal + Andaman & Nicobar Islands | Extracted | Low | 0 | 0.3 | 0.6 | 3.5 | 6.2 | 15.9 | 23.7 | 27.5 | 26.7 | 29.1 | 34.8 | 31.7 | 21.5 | 10.8 |

**Table A4. Female mortality rate of India.**
Obtained from UN life tables for 2015-2020.[15]

| Age group | India |
|---|---|
| 0-0 | 0.03290 |
| 1-4 | 0.00218 |
| 5-9 | 0.00075 |
| 10-14 | 0.00061 |
| 15-19 | 0.00100 |
| 20-24 | 0.00129 |
| 25-29 | 0.00136 |
| 30-34 | 0.00154 |
| 35-39 | 0.00201 |
| 40-44 | 0.00290 |
| 45-49 | 0.00400 |
| 50-54 | 0.00761 |
| 55-59 | 0.01060 |
| 60-64 | 0.01770 |
| 65-69 | 0.02730 |
| 70-74 | 0.04430 |
| 75-79 | 0.06650 |
| 80-84 | 0.10800 |
| 85-89 | 0.16400 |
| 90-94 | 0.24200 |
| 95-99 | 0.22500 |

**Table A5. Type-specific contribution of HPV types in cervical cancer.**
Obtained from a study of HPV distribution in cervical cancer in India.[16]

| HPV type | 16 | 18 | 31 | 33 | 35 | 39 | 45 | 52 | 56 | 58 | 59 | 68 | 73 | other | Contribution of a given combination of HPV types (%) |
|---|---|---|---|---|---|---|---|---|---|---|---|---|---|---|---|
| Combination of HPV types | x | | | | | | | | | | | | | | 57.5 |
| | | x | | | | | | | | | | | | | 10.4 |
| | | | x | | | | | | | | | | | | 1.0 |
| | | | | x | | | | | | | | | | | 3.1 |
| | | | | | | x | | | | | | | | | 0.5 |
| | | | | | | | x | | | | | | | | 1.6 |
| | | | | | | | | x | | | | | | | 1.0 |
| | | | | | | | | | x | | | | | | 1.6 |
| | | | | | | | | | | x | | | | | 1.6 |
| | | | | | | | | | | | x | | | | 2.1 |
| | x | x | | | | | | | | | | | | | 7.3 |
| | x | | x | | | | | | | | | | | | 0.5 |
| | x | | | x | | | | | | | | | | | 1.0 |
| | x | | | | x | | | | | | | | | | 1.0 |
| | x | | | | | x | | | | | | | | | 0.5 |
| | x | | | | | | | x | | | | | | | 1.0 |
| | x | | | | | | | | x | | | | | | 0.5 |
| | x | | | | | | | | | x | | | | | 0.5 |
| | x | | | | | | | | | | | x | | | 0.5 |
| | | | | | | | | | | | | | x | | 0.5 |
| | | x | | | x | | | | | | | | | | 0.5 |
| | | | | | | | | | | | | | | x | 5.8 |
| **Unnormalised contributions (%)** | 70 | 18 | 2 | 4 | 2 | 1 | 2 | 2 | 2 | 2 | 2 | 1 | 1 | 6 | |
| **Normalised contributions (%)** | 62 | 16 | 1 | 4 | 1 | 1 | 1 | 2 | 2 | 2 | 2 | 0 | 0 | 5 | |

**Table A6. Standard world population.**[18]

| Age group | Population |
|---|---|
| 0-4 | 12000 |
| 5-9 | 10000 |
| 10-14 | 9000 |
| 15-19 | 9000 |
| 20-24 | 8000 |
| 25-29 | 8000 |
| 30-34 | 6000 |
| 35-39 | 6000 |
| 40-44 | 6000 |
| 45-49 | 6000 |
| 50-54 | 5000 |
| 55-59 | 4000 |
| 60-64 | 4000 |
| 65-69 | 3000 |
| 70-74 | 2000 |
| 75-79 | 1000 |
| 80-84 | 500 |
| 85+ | 500 |
| Total | 100000 |

**Table A7. Female population size by Indian state.**
Extracted from table C-13 by the Indian Census.[19]
* States or groups of states as reported in the 2006 National Behaviour Surveillance Survey of the National AIDS Control Organization of India.[9]
ǂ Clustering of states into groups of high and low cervical cancer incidence was derived in a separate manuscript.[6]
§ Other North Eastern States include Arunachal Pradesh, Nagaland, Meghalaya, Mizoram, and Tripura.

| State/group of states * | Cluster ǂ | Population size | Percentage (%) |
|---|---|---|---|
| Andhra Pradesh | Low | 41754886 | 7.13 |
| Assam | Low | 15257203 | 2.61 |
| Bihar | Low | 49638102 | 8.48 |
| Chhattisgarh | Low | 12701295 | 2.17 |
| Delhi | High | 7793088 | 1.33 |
| Goa + Daman & Diu | Low | 811247 | 0.14 |
| Gujarat + Dadra & Nagar Haveli | Low | 28984911 | 4.95 |
| Haryana | Low | 11842082 | 2.02 |
| Himachal Pradesh | Low | 3377919 | 0.58 |
| Jammu & Kashmir | Low | 5895268 | 1.01 |
| Jharkhand | Low | 16003337 | 2.73 |
| Karnataka | High | 30108199 | 5.14 |
| Kerala + Lakshadweep | Low | 17392769 | 2.97 |
| Madhya Pradesh | High | 34975017 | 5.97 |
| Maharashtra | Low | 53942893 | 9.21 |
| Manipur | Low | 1413663 | 0.24 |
| Orissa | Low | 20704258 | 3.54 |
| Other North Eastern States § | High | 5435403 | 0.93 |
| Punjab + Chandigarh | High | 13559265 | 2.32 |
| Rajasthan | Low | 32865353 | 5.61 |
| Sikkim | Low | 286968 | 0.05 |
| Tamil Nadu + Puducherry | High | 36611821 | 6.25 |
| Uttar Pradesh | Low | 94575702 | 16.15 |
| Uttarakhand | Low | 4941223 | 0.84 |
| West Bengal + Andaman & Nicobar Islands | Low | 44595926 | 7.62 |
| Total (Low-incidence cluster) | | 456985005 | 78.05 |
| Total (High-incidence cluster) | | 128482793 | 21.94 |
| Total (all states) | | 585467798 | 100.00 |

**Table A8. Pre-vaccination risk of cervical cancer by Indian state.**

* States or groups of states as reported in the 2006 National Behaviour Surveillance Survey of the National AIDS Control Organization of India.[9]

§ Other North Eastern States include Arunachal Pradesh, Nagaland, Meghalaya, Mizoram, and Tripura.

ƚ Cases per 100,000 girls born

ǂ Cases per 100,000 woman-years

| State/group of states * | Life-time risk ƚ | Age-standardised incidence rate ǂ |
|---|---|---|
| Andhra Pradesh | 1157 | 11.5 |
| Assam | 849 | 8.9 |
| Bihar | 870 | 9 |
| Chhattisgarh | 870 | 9 |
| Delhi | 1353 | 13.7 |
| Goa + Daman & Diu | 870 | 9 |
| Gujarat + Dadra & Nagar Haveli | 868 | 9.3 |
| Haryana | 870 | 9 |
| Himachal Pradesh | 870 | 9 |
| Jammu & Kashmir | 870 | 9 |
| Jharkhand | 870 | 9 |
| Karnataka | 1667 | 16.8 |
| Kerala + Lakshadweep | 738 | 7.3 |
| Madhya Pradesh | 1498 | 15.3 |
| Maharashtra | 1158 | 11.9 |
| Manipur | 804 | 7.9 |
| Orissa | 870 | 9 |
| Other North Eastern States § | 1483 | 16.3 |
| Punjab + Chandigarh | 1304 | 13.4 |
| Rajasthan | 870 | 9 |
| Sikkim | 820 | 9.2 |
| Tamil Nadu + Puducherry | 1764 | 18.5 |
| Uttar Pradesh | 870 | 9 |
| Uttarakhand | 870 | 9 |
| West Bengal + Andaman & Nicobar Islands | 843 | 8.6 |
| Total (Low-incidence cluster) | 922 | 9.5 |
| Total (High-incidence cluster) | 1583 | 16.3 |
| Total (all states) | 1067 | 11 |

#### A.4. HPV-FRAME checklist

**Table A9. HPV-FRAME checklist [20]**
Y=yes; N=no; F=female; M=male; NA=not applicable

| **Core reporting standard** | | | |
|---|---|---|---|
| **a) Inputs** | **Reported by age? (Y/N)** | **Report by sex? (F-only, M-only or both)** | **Comments** |
| Target population for intervention | Y | Both | Routine vaccination in girls and boys were considered. Age of routine vaccination was reported. |
| Sexual behaviour | Y | Both | Parameters reports: proportion of risk-group; age- and risk-group specific numbers of stable and one-off partnerships; rates of partnership dissolution; number of sex acts per unit of time given established partnership; probability of HPV transmission per sex act. Parameters were reported separately by model for the high and low cervical cancer incidence cluster. Derivation of assortative parameters was reported. |
| Cohort examined for evaluation / time horizon | N | F-only | Life-time risk of cervical cancer was reported for the first 40 vaccinated cohorts in the routine programme. Age-standardised incidence rate of cervical cancer was reported up to 100 years after the introduction of vaccination. |
| Quality of life assumptions | NA | NA | NA |
| Calibration | Y | Y | The standard STERGM calibration function was used to obtain age- and risk-groups parameters of partnership formation and dissolution. Subsequently, maximum likelihood was used to assess the goodness-of-fit to age-specific HPV prevalence data and to obtain HPV transmission probabilities and one-off partnership underreporting rate. |
| Validation (where possible) | Y | Y | Validation adequate cervical cancer incidence resulting from the HPV infection incidence here was reported in the accompanying paper of a previous publication.[21] |
| Costs | NA | NA | NA |

| **Reporting standard for HPV vaccination in adolescent individuals** | | | | |
|---|---|---|---|---|
| **a) Inputs** | **Reported? (Y/N)** | **Reported by age? (Y/N)** | **Report by sex? (F-only, M-only or both)** | **Report as calibration or validation target? (Y/N)** |
| Vaccine uptake | Y | Y | Y | Vaccination in girls and boys were considered. Uptake between 0-100% were considered. |
| Vaccine efficacy | Y | Y | NA | Efficacy by HPV type was considered. Efficacy was independent of age or gender. |
| Vaccine cross-protection | Y | Y | NA | Level of cross-protection for HPV 31/33/45 was reported. |
| Duration vaccine protection and waning | Y | Y | NA | Lifelong vaccine protection was considered based on the evidence from the IARC India vaccine trial.[22] |
| Vaccine and delivery costs | NA | NA | NA | Economic assessment was not performed. |
| Pre-vaccination disease burden (including population attributable fractions for HPV) | Y, for cervical cancer. | Y, for cervical cancer. | F-only, for cervical cancer. | Attributable fraction by HPV type to cervical cancer burden was reported. |
| Duration of natural immunity | Y | NA | Y | Natural immunity was independent of age. Sex-specific natural immunity assumptions were reported. |
| **b) Outputs** | **Reported? (Y/N)** | **Reported by age? (Y/N)** | **Report by sex? (F-only, M-only or both)** | **Comments** |
| Absolute reductions in HPV infections, and/or warts, post-vaccination | N | NA | NA | Impact on HPV prevalence and warts were not considered. |
| Absolute reductions in CIN2+ post-vaccination | N | NA | NA | NA |

(Table continues the next page)

| | | | | |
|---|---|---|---|---|
| Absolute reductions in invasive cancer (cervical and other HPV cancers, as relevant) | Y, for cervical cancer. | N | F-only, cervical cancer. | NA |

| Reporting standard for models of HPV prevention in LMIC | | | | |
|---|---|---|---|---|
| **a) Inputs** | **Reported? (Y/N)** | **Reported by age? (Y/N)** | **Report by sex? (F-only, M-only or both)** | **Comments** |
| HIV prevalence rates if endemic in country | N | N | N | The effects of HIV are not modelled because of the low HIV prevalence in India.[3] |
| Description of any opportunistic or pilot/demonstration screening project ongoing | Y | NA | NA | Opportunistic cervical cancer screening is done in the Indian national communicable disease control programme. However, the coverage is very low. Ever-in-lifetime coverage was reported to be lower than 3% for women aged 30–49 years in the 2017–18 NCD monitoring survey.[23] Hence, no screening was assumed in the present study. |

| Reporting standards for evaluations assessing alternative vaccine types or reduced-dose schedules | | | | |
|---|---|---|---|---|
| **a) Inputs** | **Reported? (Y/N)** | **Reported by age? (Y/N)** | **Report by sex? (F-only, M-only or both)** | **Comments** |
| Vaccine efficacy/waning | Y | See Comments. | See Comments. | Efficacy by HPV type was considered. Efficacy was independent of age or gender. Lifelong vaccine protection was considered based on the evidence from the IARC India vaccine trial representing single-, two-, or three-dose scehdules.[22] |
| Timing between doses (for 2-dose) | NA | NA | NA | Timing between the two doses under two-dose vaccination schedule was not modelled. We assumed constant and life-long efficacy. |
| Vaccine cross-protection | Y | See Comments. | See Comments. | Level of cross-protection for HPV 31/33/45 was reported. Cross-protection was independent of age or gender. |
| Cost | NA | NA | NA | NA |
| **b) Outputs** | **Reported? (Y/N)** | **Reported by age? (Y/N)** | **Report by sex? (F-only, M-only or both)** | **Report as calibration or validation target (Y/N)?** |
| Threshold cost per dose | NA | NA | NA | NA |

### Appendix B

#### B.1. Supplementary results in tables

**Table B1. Sensitivity analyses on coverage at disruption and duration of disruption on resilience by Indian state.** Life-time number of cervical cancer cases prevented per 100,000 girls born in birth cohorts vaccinated prior to disruption in part I. Sensitivity analyses on coverage at disruption in part II and on duration of disruption in part III on resilience (defined as the life-time number of cervical cancer cases still prevented in the birth cohorts with disruption of vaccination per 100,000 girls born) and resilience ratio (defined as fold change in resilience by switching from one scenario to another). Uncertainty intervals are reported in brackets.

*I. Life-time number of cervical cancer cases prevented prior to disruption*

| Scenario | States | GO 60% | GO 90% | GN 60% | GN 90% |
|---|---|---|---|---|---|
| No disruption | All | 562 (444, 676) | 773 (701, 836) | 647 (539, 746) | 807 (752, 853) |
| | High | 852 (706, 995) | 1149 (1062, 1223) | 936 (777, 1081) | 1197 (1130, 1260) |
| | Low | 481 (370, 586) | 668 (599, 727) | 566 (472, 651) | 697 (646, 739) |

*II. Sensitivity analyses on coverage at disruption (with duration of disruption fixed at 5 years)*

| Coverage at disruption in % | States | Resilience by vaccination strategy and coverage | | | | Resilience ratio | | |
|---|---|---|---|---|---|---|---|---|
| | | GO 60% | GO 90% | GN 60% | GN 90% | GO 60% to GO 90% | GO 60% to GN 60% | GO 90% to GN 90% |
| 0 (base case) | All | 107 (7, 214) | 209 (81, 340) | 302 (170, 437) | 464 (328, 602) | 2.0 | 2.8 | 2.2 |
| | High | 151 (33, 281) | 291 (136, 457) | 459 (291, 628) | 693 (515, 876) | 1.9 | 3.0 | 2.4 |
| | Low | 95 (0, 195) | 186 (66, 307) | 257 (136, 383) | 399 (276, 525) | 2.0 | 2.7 | 2.2 |
| 20 | All | 271 (155, 391) | 355 (221, 490) | 425 (297, 559) | 550 (416, 680) | 1.3 | 1.6 | 1.6 |
| | High | 393 (235, 555) | 508 (341, 678) | 638 (460, 823) | 820 (647, 989) | 1.3 | 1.6 | 1.6 |
| | Low | 237 (132, 345) | 312 (187, 438) | 366 (251, 485) | 474 (351, 593) | 1.3 | 1.5 | 1.5 |
| 40 | All | 410 (277, 534) | 476 (343, 599) | 527 (401, 647) | 621 (500, 730) | 1.2 | 1.3 | 1.3 |
| | High | 603 (442, 757) | 697 (533, 853) | 788 (631, 929) | 926 (777, 1060) | 1.2 | 1.3 | 1.3 |
| | Low | 356 (231, 471) | 414 (289, 527) | 453 (337, 567) | 536 (423, 638) | 1.2 | 1.3 | 1.3 |

*III. Sensitivity analyses on duration of disruption (with coverage at disruption fixed at 0%)*

| Duration of disruption in years | States | Resilience by vaccination strategy and coverage | | | | Resilience ratio | | |
|---|---|---|---|---|---|---|---|---|
| | | GO 60% | GO 90% | GN 60% | GN 90% | GO 60% to GO 90% | GO 60% to GN 60% | GO 90% to GN 90% |
| 1 | All | 137 (26, 253) | 261 (125, 407) | 365 (215, 502) | 517 (372, 655) | 1.9 | 2.7 | 2.0 |
| | High | 202 (63, 345) | 372 (183, 575) | 549 (350, 726) | 767 (605, 930) | 1.8 | 2.7 | 2.1 |
| | Low | 119 (16, 227) | 230 (109, 359) | 313 (177, 439) | 447 (307, 578) | 1.9 | 2.6 | 1.9 |
| 2 | All | 125 (17, 233) | 240 (105, 375) | 344 (206, 480) | 500 (359, 642) | 1.9 | 2.7 | 2.1 |
| | High | 183 (54, 324) | 338 (167, 519) | 507 (333, 692) | 738 (553, 929) | 1.9 | 2.8 | 2.2 |
| | Low | 109 (6, 207) | 212 (87, 334) | 298 (171, 421) | 433 (305, 561) | 2.0 | 2.7 | 2.0 |
| 5 (base case) | All | 107 (7, 214) | 209 (81, 340) | 302 (170, 437) | 464 (328, 602) | 2.0 | 2.8 | 2.2 |
| | High | 151 (33, 281) | 291 (136, 457) | 459 (291, 628) | 693 (515, 876) | 1.9 | 3.0 | 2.4 |
| | Low | 95 (0, 195) | 186 (66, 307) | 257 (136, 383) | 399 (276, 525) | 2.0 | 2.7 | 2.2 |
| 10 | All | 80 (0, 182) | 154 (33, 275) | 226 (96, 358) | 382 (240, 525) | 1.9 | 2.8 | 2.5 |
| | High | 113 (0, 241) | 217 (64, 367) | 340 (168, 506) | 560 (381, 741) | 1.9 | 3.0 | 2.6 |
| | Low | 71 (0, 165) | 137 (25, 249) | 194 (76, 317) | 332 (200, 464) | 1.9 | 2.8 | 2.4 |

#### B.2. Supplementary results in figures

**Figure B1. Resilience against HPV vaccination disruption in the base case by Indian state.** Predicted HPV vaccination resilience, defined as life-time number of cervical cancer cases still prevented in the birth cohorts with disruption of vaccination per 100,000 girls born (blue arrow), and drop in cervical cancers prevented as compared to impact in the undisrupted cohorts (black arrow), under the four highlighted scenarios in Indian states with (A) high and (B) low cervical cancer incidence. Disruption was simulated according to the base case with a period of disruption of 5 years and 0% coverage in girls and boys during the disruption period. Figure 1 in the main text corresponds to the results for all Indian states.

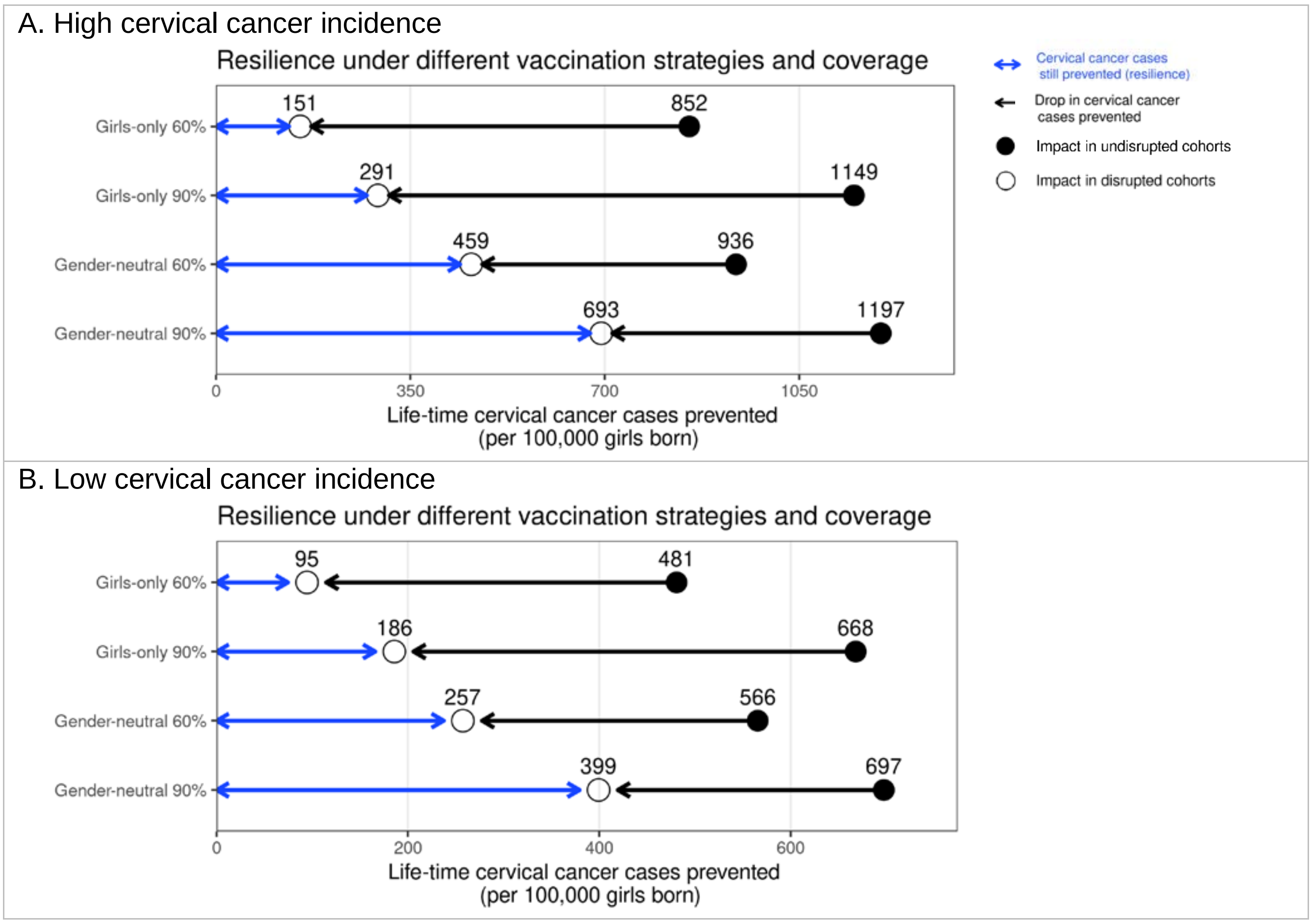

**Figure B2. Resilience against HPV vaccination disruption in sensitivity analyses on coverage at disruption by Indian state.** Predicted HPV vaccination resilience, defined as life-time number of cervical cancer cases still prevented in the birth cohorts with disruption of vaccination per 100,000 girls born (blue arrow), and drop in cervical cancers prevented as compared to impact in the undisrupted cohorts (black arrow), under different vaccination strategies and coverage (rows) in the sensitivity analyses of the coverage of disruption in Indian states with (A) high and (B) low cervical cancer incidence. Disruption was simulated for 0% (base-case), 20%, or 40% coverage in girls at disruption. The vaccination coverage was fixed at 0% in boys, and the duration of disruption was fixed at 5 years.

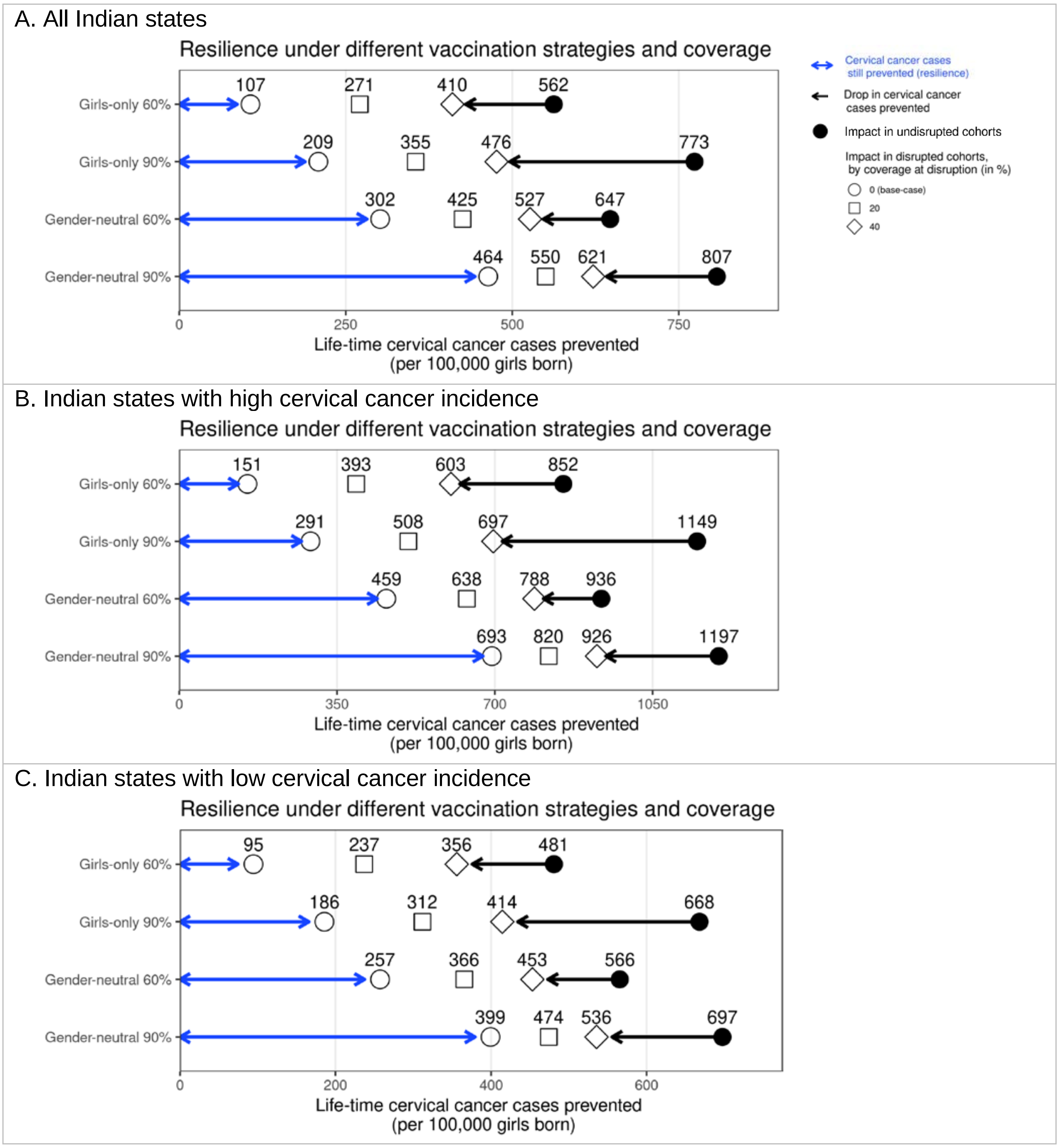

**Figure B3. Resilience against HPV vaccination disruption in sensitivity analyses on duration of disruption by Indian state.** Predicted HPV vaccination resilience, defined as life-time number of cervical cancer cases still prevented in the birth cohorts with disruption of vaccination per 100,000 girls born (blue arrow), and drop in cervical cancers prevented as compared to impact in the undisrupted cohorts (black arrow), under different vaccination strategies and coverage (rows) in the sensitivity analyses of the duration of disruption in (A) all Indian states, (B) Indian states with high cervical cancer incidence and (C) Indian states with low cervical cancer incidence. Disruption was simulated for 1, 5 (base-case), or 10 years indicated by different empty shapes. Vaccination coverage was fixed at 0% in girls and boys during the period of disruption.

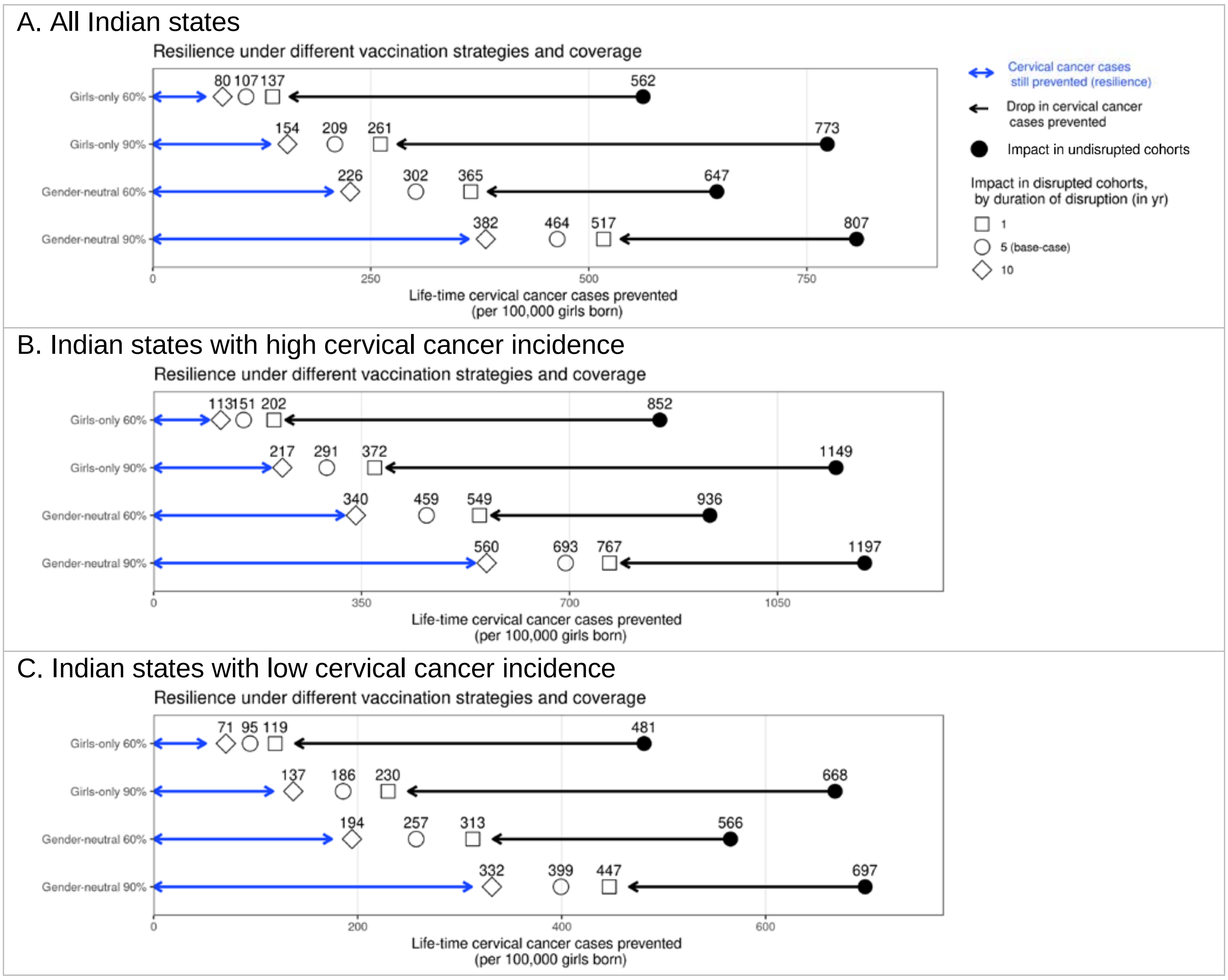

**Figure B4. Progress towards cervical cancer elimination over time with and without disruption.** Predicted cervical cancer age-standardised incidence (in cases per 100,000 woman-years) in the years since start of vaccination in the four highlighted scenarios with (dashed curves) and without (solid curves) disruption. The dashed horizontal line represents the WHO elimination threshold for cervical cancer elimination, i.e., age-standardised incidence of 4 cases per 100,000 woman-years. Disruption was simulated according to the base case with a period of disruption of 5 years and 0% coverage in girls and boys during the disruption period.

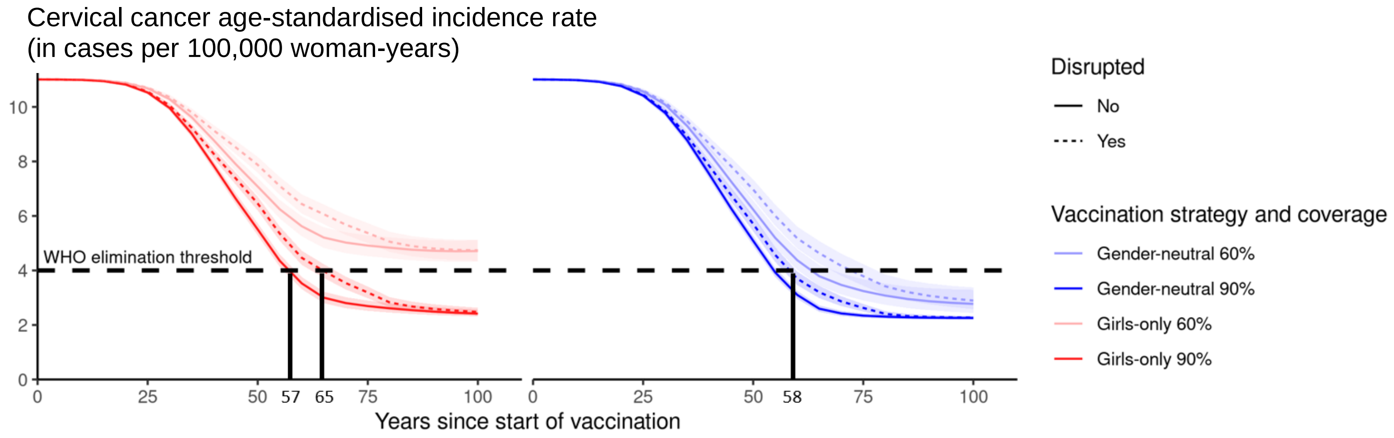
